# Serological evaluation of a cluster randomised trial on the use of reactive focal mass drug administration and reactive vector control to reduce malaria transmission in Zambezi Region, Namibia

**DOI:** 10.1101/2021.04.12.21255334

**Authors:** Lindsey Wu, Michelle S. Hsiang, Lisa M. Prach, Leah Schrubbe, Henry Ntuku, Mi-Suk Kang Dufour, Brooke Whittemore, Valerie Scott, Joy Yala, Kathryn W. Roberts, Catriona Patterson, Joseph Biggs, Tom Hall, Kevin K.A. Tetteh, Cara Smith Gueye, Bryan Greenhouse, Adam Bennett, Jennifer L. Smith, Stark Katokele, Petrina Uusiku, Davis Mumbengegwi, Roly Gosling, Chris Drakeley, Immo Kleinschmidt

## Abstract

Due to challenges in measuring changes in malaria in low transmission settings, serology is increasingly being used to complement clinical and parasitological surveillance. Longitudinal cohort studies have shown serological markers, such as Etramp5.Ag1, to be particularly discriminatory of spatio-temporal differences in malaria transmission. However, these markers have yet to be used as endpoints in intervention trials. This study is an extended analysis of a 2017 cluster randomised trial conducted in Zambezi Region, Namibia, evaluating the effectiveness of reactive focal mass drug administration (rfMDA) and reactive vector control (RAVC). A panel of eight serological markers of *Plasmodium falciparum* infection - Etramp5.Ag1, GEXP18, HSP40.Ag1, Rh2.2030, EBA175, *Pf*MSP1_19_, *Pf*AMA1, and *Pf*GLURP.R2 - was used on a multiplex immunoassay to measure population antibody responses as trial endpoints.

Reductions in sero-prevalence to antigens Etramp.Ag1, *Pf*MSP1_19_, Rh2.2030, and *Pf*AMA1 were observed in study arms combining rfMDA and RAVC, but only effects for Etramp5.Ag1 were statistically significant. Etramp5.Ag1 sero-prevalence was significantly lower in all intervention arms. Compared to the reference arms, adjusted Etramp5.Ag1 prevalence ratio (aPR) was 0.77 (95%CI 0.65 – 0.90, p<0.001) for rfMDA and 0.79 (95%CI 0.67 – 0.92, p=0.001) for RACD. For combined rfMDA plus RAVC, aPR was 0.58 (95%CI 0.45 – 0.75, p<0.001). Significant reductions were also observed based on continuous antibody responses. Sero-prevalence as an endpoint was found to achieve higher study power (99.9% power to detect a 50% reduction in prevalence) compared to quantitative polymerase chain reaction (qPCR) prevalence (72.9% power to detect a 50% reduction in prevalence).

The use of serological endpoints to evaluate trial outcomes was comparable to qPCR and measured effect size with improved precision. Serology has clear application in cluster randomised trials, particularly in settings where measuring clinical incidence or infection is less reliable due to seasonal fluctuations, limitations in health care seeking, or incomplete testing and reporting.

**Key questions:** *What is already known?:*

▪ Numerous serological studies across sub-Saharan Africa have found that malaria-specific antibody responses are highly correlated with malaria transmission.
▪ Serology is increasingly being used to complement traditional malaria surveillance data in settings where clinical or parasitological measures of incidence or infection may be less reliable due to fluctuations in parasite densities, limitations in health care seeking, or incomplete testing and reporting.
▪ The identification of new serological markers associated with recent malaria exposure hold promise as measures of malaria incidence. In previous longitudinal cohort studies, Etramp5.Ag1 has been shown to be a discriminatory serological marker capable of detecting spatio-temporal differences in malaria transmission. However, these markers have never been formally used as endpoints in a malaria cluster randomised trial.

*What are the new findings?:*

▪ This study is the first application of serological endpoints in a malaria cluster randomised trial. Using a multiplexed immunoassay, a panel of sero-incidence markers of recent malaria exposure were used to evaluate the effectiveness of reactive focal mass drug administration (rfMDA) and reactive focal vector control (RAVC) compared to reactive case detection (standard of care) to reduce malaria transmission.
▪ Cluster-level antibody responses were significantly lower in all intervention arms compared to control, and effect sizes were measured with greater study power than other trial endpoints such as quantitative polymerase chain reaction (qPCR) parasite prevalence.

*What do the new findings imply?:*

▪ The findings from this study, together with ongoing innovations in assay design and multi-disease platforms, illustrate the potential application of serological markers as endpoints in cluster randomised trials. The use of serological endpoints can help achieve trial efficiencies, such as reduced sample size, particularly in low transmission settings or multi-intervention trials where measuring differences between study arms may be challenging with clinical or parasitological endpoints alone.

## Introduction

Namibia is one of a number of southern African countries targeting malaria elimination, and, in 2009, the Elimination 8 initiative was created to support these goals region-wide.^1,2^ Between 2001 and 2011, Namibia experienced a marked epidemiologic transition, during which clinical cases of malaria fell by 97.4% and malaria-attributable deaths by 98%.^3^ This was the result of policies for indoor-residual spraying (IRS) in endemic areas, improved case management with rapid diagnostic tests (RDTs), and access to artemisinin combination therapies (ACTs).^3–6^ However, recent progress towards malaria elimination has stalled. Despite decades of widespread IRS and the introduction of reactive case detection (RACD) in 2012, areas of northern Namibia experienced significant outbreaks during the 2016 malaria season,^7^ and these periodic spikes in incidence have created unexpected challenges for the planning of elimination efforts.

RACD is widely used for actively targeting asymptomatically infected residents in the neighbourhood of passively identified index cases,^8^ but the limited sensitivity of current field diagnostics^9–11^ may be one reason RACD has not demonstrated effectiveness at low transmission.^12–14^ To accelerate malaria elimination in settings such as Namibia, new focal approaches such as reactive focal mass drug administration (rfMDA) and reactive vector control (RAVC) have been suggested, whereby MDA or vector control is targeted at the infectious human or mosquito reservoir specifically in high-risk areas in close proximity to a passively-detected index case. In 2017, a cluster randomised control trial was conducted in Zambezi Region, Namibia, evaluating rfMDA and RAVC compared to RACD as standard of care and observed a reduction in clinical incidence and prevalence of infection measured by quantitative polymerase chain reaction (qPCR).^15^

In elimination settings, measuring changes in transmission can be imprecise due to low levels of clinical incidence, making adequate sample sizes difficult to achieve. As a result, serological methods are increasingly being used alongside clinical and parasitological metrics to complement traditional surveillance data. Studies in moderate to low transmission regions across sub-Saharan Africa, including Tanzania^16^, Equatorial Guinea^17^, South Africa^18^ and The Gambia^19^, have found that malaria-specific antibody responses are highly correlated with parasitological endpoints. Serological assays can be a particularly cost-effective tool in settings where parasite densities commonly fall below the detection limit of other field diagnostics such as RDTs. In some areas, serology has also been used to confirm interruption of malaria transmission.^20,21^

Most serological studies for malaria have monitored historical trends based on long-lived antibody responses rather than with sero-incidence markers associated with short-lived antibody responses. Sero-incidence markers aim to measure recent exposure or detect rapid changes in transmission (e.g., over periods of 1-5 years). Recent studies analysing cohorts in Uganda and Mali have identified several serological markers that are predictive of clinical malaria in the previous year^22^. Etramp5.Ag1 in particular has been shown to be a discriminatory sero-incidence measure between geographical regions and transmission seasons in The Gambia^19,23^. However, serological markers of malaria exposure have rarely been used as outcome measures for cluster randomised trials.

The study presented here is an extended trial analysis assessing cluster-level antibody responses to a panel of serological markers previously shown to be associated with parasitological measures of malaria.^19,23^ Using a multiplexed bead-based assay and samples from the endline cross-sectional survey of the RACD/rfMDA/RAVC trial, the aim of this study was to evaluate the trial interventions using serological markers of recent malaria infection. These are compared with the primary trial endpoints of malaria case incidence and parasite infection prevalence. Additionally, trial design parameters, including inter-cluster coefficient of variation and trial sample sizes, were re-estimated to assess whether trial efficiencies can be achieved using serological endpoints.

## Materials and Methods

### Data and study design

The study was an open label cluster randomised controlled trial with a 2×2 factorial study design (Supplementary Figure S1) with four study arms receiving the following interventions:

1. **RACD (standard of care control arm):** rapid diagnostic testing and treatment of positives with artemether-lumefantrine (AL) and single dose primaquine of individuals residing within a 500m radius of a recent passively detected index case
2. **rfMDA:** presumptive treatment with artemether-lumefantrine (AL) of individuals residing within a 500m radius of a recent passively detected index case
3. **RAVC and RACD combined:** indoor residual spraying (IRS) using pirimiphos-methyl, administered to households of individuals residing within a 500m radius of a recent passively detected index case, plus standard of care RACD as described above
4. **rfMDA and RAVC combined:** indoor residual spraying (IRS) using pirimiphos-methyl, administered to households of individuals residing within a 500m radius of a recent passively detected index case, plus rfMDA as described above

All clusters received routine annual IRS before the start of the malaria season using dichloro-diphenyl-trichloroethane (DDT) conducted as part of standard malaria control activities by the Namibian Ministry of Health and Social Services (MoHSS).

The trial was conducted in Zambezi Region, Namibia, from January to November 2017, within the catchment areas for 11 health facilities. Of the 102 enumeration areas (EAs) in the study area, 46 clusters met the exclusion criteria (no incident cases of malaria between 2012 and 2014 or incomplete incidence data due to missing records), and the remaining 56 clusters were selected and randomly allocated to one of four arms using restricted randomisation. Restriction criteria included mean annual incidence in 2013 and 2014, population size, population density, and mean distance from the household to a health-care facility. The primary outcome of the main study was cumulative incidence of passively detected malaria. Secondary outcomes included infection prevalence, intervention coverage, refusal rates, adverse events, and adherence to drug regimen. Details of the study are reported on ClinicalTrials.gov: NCT02610400^24^ and described in Medzihradsky et al, 2018.^25^ The clinical and parasitological results of the trial are reported in Hsiang et al.^15^

An endline cross-sectional survey was conducted as part of the trial at the end of the malaria season from May to August 2017 to measure infection prevalence by qPCR and sero-prevalence. Within each of the 56 clusters, 25 households were randomly sampled for inclusion in the cross-sectional survey. All residents older than six months who slept in the household at least three nights per week in the previous four weeks were eligible for inclusion in the cross-sectional survey. For consenting individuals, blood samples were collected by finger prick for molecular and serological analysis on dry blood spot (DBS) filter paper (Whatman 3 Corporation, Florham Park, NJ, USA) and 250 μl of whole blood in BD Microtainer® tubes with EDTA additive (Becton, Dickinson and Corporation, Franklin Lakes, NJ, USA). Individuals, both symptomatic and asymptomatic, with positive RDT results were treated with AL and single dose primaquine according to national guidelines.^26^

The trial received ethical approval from the Namibia MoHSS (17/3/3), and the Institutional Review Boards of the University of Namibia (MRC/259/2017), University of California San Francisco (15– 17422) and London School of Hygiene & Tropical Medicine (10411). Study findings were shared in stakeholder meetings attended by regional and national MoHSS representatives, with health facility staff and community leaders, through peer-reviewed publications and at scientific conferences.

### Laboratory procedures

Human plasma from whole blood samples were prepared and tested on the Luminex assay platform using procedures described by Wu et al.^27^ Plasma samples from study participants were prepared from 250 μl of whole blood collected in BD Microtainer tubes with EDTA additive in a 1:200 sample dilution. Two sets of positive controls were used based on pooled sera from 100 hyper-immune Tanzanian individuals and a WHO malaria reference lyophilised serum reagent (NIBSC 10-198).^28^ Plasma samples from European malaria-naïve adults (1:200 dilution) were used as negative controls. Two wells on each plate containing only antigen-coupled beads and sample buffer were included to measure background signal.

#### Antigen selection and design

A subset of eight antigens (Etramp5.Ag1, GEXP18, HSP40.Ag1, Rh2.2030, EBA175, *Pf*MSP1_19_, *Pf*AMA1, *Pf*GLURP.R2) were selected from an initial screen of 856 candidates on an *in vitro* transcription and translation (IVTT) protein microarray based on their correlation with clinical and parasitological endpoints in previous studies.^19,23^ Antigens were expressed in *Escherichia coli* (*E*.*coli*) as glutathione S-transferase (GST)-tagged fusion proteins, except for *Pf*AMA1 expressed in Pichia pastoris as a histidine-tagged protein. Non-malaria reactivity against GST-tagged fusion proteins were assessed using IgG responses to GST-coupled beads, and samples with greater than 1000 median fluorescence intensity (MFI) were excluded from analyses due to high non-specific IgG response. A full description of laboratory methods, including antigen constructs, expression platform, coupling conditions and data standardisation are detailed further in Wu et al^27^ and summarised in Supplementary Table S2. Antigens were classified as short-, medium-, and long-term markers of malaria exposure based on the magnitude of the association of antibody responses with age as a proxy for longevity of antibody response (Supplementary Table S3).

### Statistical analyses

Serological responses were used as endpoints to assess the following effects (Supplementary Figure S1):

1. rfMDA vs. RACD (with or without RAVC)
2. RAVC vs No RAVC (with either RACD or rfMDA)
3. rfMDA plus RAVC vs. RACD only

In line with the primary trial analysis, reported by Hsiang et al^15^, a modified intention-to-treat analysis was conducted, which adjusted at the EA-level for baseline incidence in 2016, intervention response time, proximity to a Namibia MoHSS co-intervention, index case coverage (the proportion of eligible index cases covered by an intervention), and the household coverage of the target population (the proportion of eligible individuals or households within an intervention event area triggered by an index case that actually received the intervention). Proximity to a MoHSS co-intervention was defined as households within 500m of a village that received concomitant additional interventions carried out by the Namibia MoHSS in the form of active case detection or IRS. As these additional interventions in close proximity to some of the study clusters could not be balanced between study arms, it was necessary to adjust for this as a variable in the analysis. For RACD and rfMDA study arms, the index case coverage was defined as the proportion of index cases where the intervention was implemented in households covering at least 25 individuals within 5 weeks of case identification. For the RAVC study arms, index case coverage was defined as the proportion of index cases where at least 7 households were sprayed within 5 weeks of case identification.

Serological responses were measured as median fluorescence intensity (MFI) values, normalised to account for between plate variation. Sero-positivity values for all samples were assigned according to MFI thresholds defined by the mean and standard deviation of a pool of 71 malaria naïve blood donors used as negative controls.^19,27^ For antigens associated with longer-lived antibody responses (*Pf*MSP1_19_, *Pf*AMA1, and *Pf*GLURP.R2), individuals previously exposed but not infected for 5 or more years may still have higher average antibody responses^29,30^ than malaria-naïve donors and represent a more accurate population to define endemic sero-negative thresholds. For these markers, sero-positivity MFI thresholds were defined as the mean and standard deviation of the negative component of a Gaussian mixture model fit to antibody responses of the endemic population. Serological responses were then assessed to compare the intervention study arms.

### Sero-prevalence

Sero-prevalence of population antibody responses to each antigen was estimated using generalised linear models (GLM) with log link, binomial family, generalised estimating equations (GEE) allowing for within cluster correlation and assessed according to the three interventions described above. Results are presented as unadjusted mean sero-prevalence by intervention and study arm and as unadjusted and adjusted sero-prevalence ratios, where the denominator is mean sero-prevalence of clusters in the reference study arm (RACD only). Additionally, the combined sero-prevalence to any short-term marker (i.e. sero-positivity to at least one of Etramp5.Ag1, GEXP18 or HSP40.Ag1) as an overall measure of sero-incidence was also estimated to assess whether positivity to multiple endpoints would result in an enhanced comparison between interventions.

### Antibody acquisition

Dichotomisation of data into positive and negative categories, the basis for estimating sero-prevalence, can lead to some loss of sensitivity in detecting changes in malaria transmission.^17^ Therefore, changes in the magnitude of antibody responses were also assessed using antibody acquisition models, which estimates the geometric mean MFI by age^23^ for each antigen and cluster. Using this model fit, the total area under the antibody acquisition curve, referred to in this analysis as the AUC value, represents the cumulative antibody response across all ages. The effect of the intervention on mean AUC across clusters was assessed using GLM (with log link and gaussian family), inverse-weighted by the 95% CI of the cluster AUC values, and results are presented as unadjusted and adjusted AUC ratios, where the denominator is mean AUC value of clusters in the reference study arm.

For both sero-prevalence and antibody acquisition, regression analysis tested the effect of each intervention independently as well as with an interaction between rfMDA and RAVC (Supplementary Tables S5 and S6), allowing assessment of the effect of each intervention singly (rfMDA or RAVC) or in combination (rfMDA plus RAVC). This allows interpretation of whether the combination of rfMDA and RAVC interventions were simply additive, synergistic, or antagonistic

### Between cluster coefficient of variation and sample size calculations

To assess how trial sample sizes might be affected by using serological outcome measures for the design of a study, we estimated the between cluster coefficient of variation, k,^31^ for sero-prevalence to Etramp5.Ag1 (the best performing individual marker) to compare these to corresponding values of k for qPCR. Analysis was limited to RACD only arms as a proxy for baseline antibody patterns. The required number of clusters per arm, c,^31^ was calculated to demonstrate a change in sero-prevalence of 50% in each of the RAVC or rfMDA arms and 75% in the the rfMDA plus RAVC arm, compared to RACD only as the control. These percentages were the expected reductions in clinical incidence upon which the main study was designed. Sample size (number of clusters per study arm) was estimated for a range of cluster sizes, assuming a desired study power of 80% and a 5% two-sided significance level. As a comparator, the coefficient of variation and minimum sample sizes were also calculated using qPCR prevalence as the endpoint.

## Results

### Study participant characteristics and intervention implementation and coverage

A total of 4,361 individuals were enrolled in the end-line cross-sectional survey, of which 4,164 samples were available for serological processing. After excluding samples with high GST values to avoid the influence of non-malaria specific antibody response, a total of 3,657 samples were available for final analysis. Age and gender distribution were similar across study arms (Supplementary Table S1). Less than 10% of participants reported sleeping outdoors in the previous two weeks and only a quarter of participants reported sleeping under a bed net the previous night. Intervention coverage by index case and target population and proximity to MoHSS co-intervention were reported previously.^15^ Briefly, coverage ranged from 87% to 95.2% of the target population, and between 43.4% and 61.8% of households (by study arm) were within 500m of a village that received concomitant additional interventions carried out by the Namibia MoHSS.

### Sero-prevalence by study arm

Population antibody responses varied by antigen, with sero-prevalence in the RACD only arm ranging from 0.22 (95%CI 0.20 – 0.27) for HSP40.Ag1 and 0.28 (95%CI 0.24 – 0.33) for Etramp5.Ag1 to 0.48 (95%CI 0.45 – 0.52) for *Pf*AMA1 (Supplementary Figure S2, Table S4). The largest reductions in sero-prevalence were observed in the rfMDA plus RAVC compared to the RACD only arms for Etramp5.Ag1, *Pf*MSP1_19_, Rh2.2030, and *Pf*AMA1 (Figure 1A). However, results were only statistically significant for antibody responses to Etramp5.Ag1.

**Figure 1.**
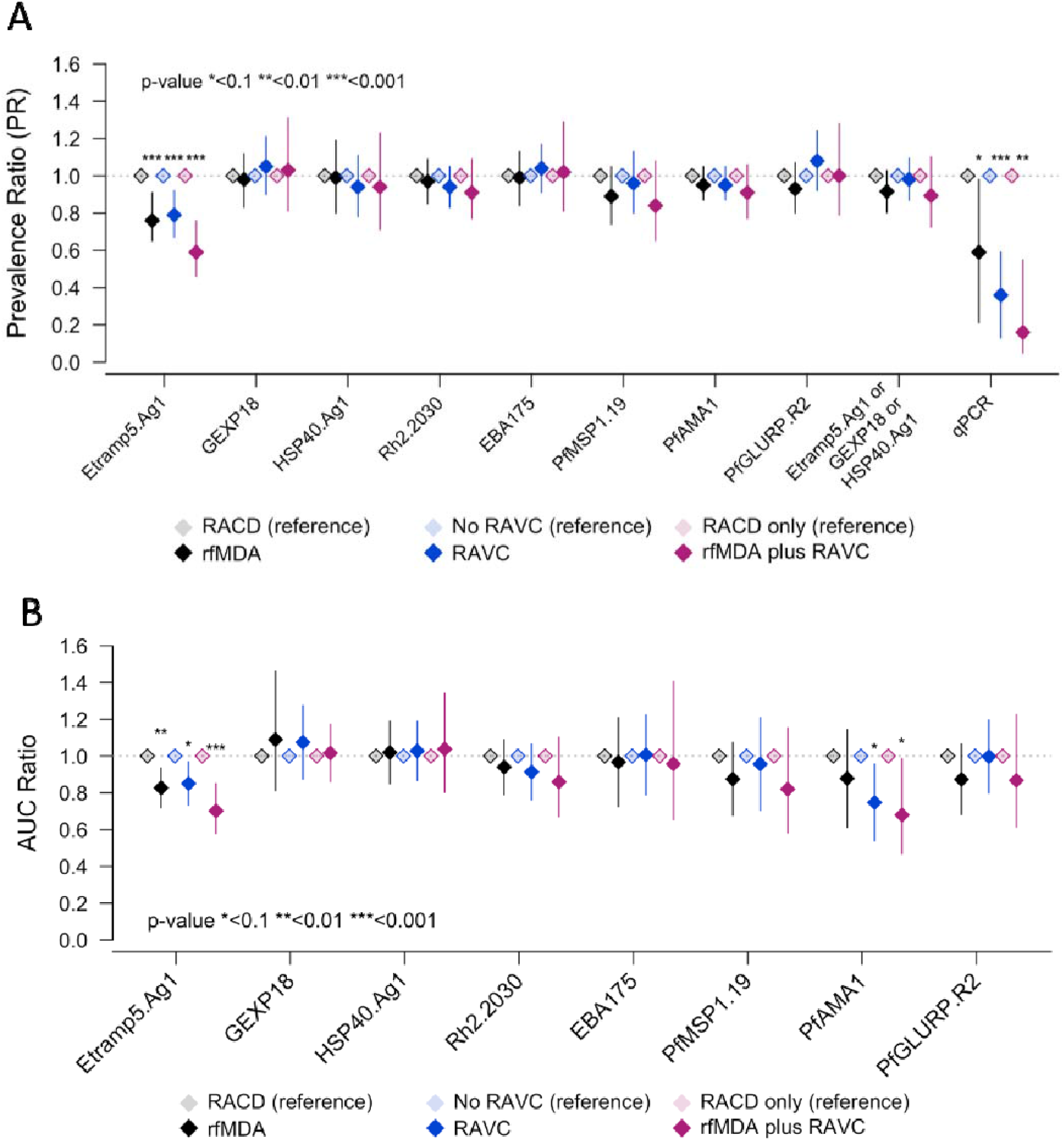
Sero-prevalence ratio, qPCR prevalence ratio, and AUC ratio by antigen and intervention. Adjusted prevalence ratios are shown for rfMDA vs RACD (black), RAVC vs. no RAVC (blue) and rfMDA plus RAVC vs. RACD only (magenta). Adjusted ratio of log AUC values are shown for rfMDA vs RACD (black), RAVC vs. no RAVC (blue) and rfMDA plus RAVC vs. RACD only (magenta). All values are adjusted for EA incidence in 2016, proportion of EA index cases covered, proportion of target population covered, median time to intervention, distance from villages receiving an MOHSS intervention.

Sero-prevalence to Etramp5.Ag1 was significantly lower in all intervention arms compared to the reference arms (Figure 1A, Table 1). In the rfMDA arms, unadjusted mean Etramp5.Ag1 sero-prevalence was 0.22 (95% CI 0.19 – 0.25) compared to 0.28 (95%CI 0.25 – 0.30) in the RACD reference arms, adjusted prevalence ratio (aPR) 0.78 (95%CI 0.65 – 0.91, p<0.001). Mean Etramp5.Ag1 sero-prevalence for RAVC arms was 0.23 (95%CI 0.20 – 0.26) vs 0.27 (95%CI 0.24 – 0.31) in the non-RAVC arms, aPR 0.79 (95%CI 0.67 – 0.92, p=0.001). The largest effect was observed in the rfMDA plus RAVC arms, where mean Etramp5.Ag1 sero-prevalence was 0.18 (95%CI 0.14 – 0.25) compared to 0.28 (95%CI 0.24 – 0.33) in the RACD only arms, aPR 0.59 (95%CI 0.46 – 0.76, p<0.001).

**Table 1.** Etramp5.Ag1 sero-prevalence ratio by intervention. Mean sero-prevalence and sero-prevalence ratios are estimated using generalised linear models by intervention (with log link, binomial family, and GEE with clustering at EA-level). Prevalence ratios are adjusted for EA incidence in 2016, proportion of EA index cases covered, proportion of target population covered, median time to intervention, and distance from villages receiving an MOHSS intervention.

|  |  |  | Unadjusted |  |  | Adjusted |  |
| --- | --- | --- | --- | --- | --- | --- | --- |
|  | Number of individuals | Number of clusters | Mean prevalence (95% CI) | Prevalence ratio (95% CI) | p-value | Prevalence ratio (95% CI) | p-value |
| **Human reservoir** |  |  |  |  |  |  |  |
| RACD (reference)* | 1,993 | 27 | 0.28 (0.25 – 0.31) | 1.00 | -- | 1.00 | -- |
| rfMDA* | 1,664 | 28 | 0.22 (0.19 – 0.25) | 0.78 (0.64 – 0.93) | 0.004 | 0.78 (0.65 – 0.91) | <0.001 |
| **Mosquito reservoir** |  |  |  |  |  |  |  |
| No RAVC (reference)† | 1,905 | 27 | 0.27 (0.24 – 0.31) | 1.00 | -- | 1.00 | -- |
| RAVC† | 1,752 | 28 | 0.23 (0.20 – 0.26) | 0.85 (0.70 – 1.00) | 0.057 | 0.79 (0.67 – 0.92) | 0.001 |
| **Human and mosquito reservoir** |  |  |  |  |  |  |  |
| RACD only (reference) | 990 | 13 | 0.28 (0.24 – 0.33) | 1.00 | -- | 1.00 | -- |
| rfMDA plus RAVC | 749 | 14 | 0.18 (0.14 – 0.25) | 0.65 (0.49 – 0.87) | 0.003 | 0.59 (0.46 – 0.76) | <0.001 |

\* With or without RAVC † With either RACD or rfMDA

Similar effects were observed in the main study, as shown in Figure 1A, where the adjusted qPCR prevalence ratio was 0.59 (95%CI 0.21 – 0.98, p=0.039) for rfMDA versus RACD, 0.36 (95%CI 0.13 – 0.59, p<0.0001) for RAVC versus non-RAVC, and 0.16 (95%CI 0.05 – 0.55, p=0.004) for rfMDA plus RAVC versus RACD only. Sero-prevalence to a combination of sero-incidence markers (Etramp5.Ag1, GEXP18, and HSP40.Ag1) were also assessed, but no significant differences between study arms were observed. The distribution of cluster-level sero-prevalence by study arm is shown in Supplementary Figure S4.

### Antibody acquisition by study arm

Analysis of cluster-level Etramp5.Ag1 AUC values based on continuous antibody response confirmed smaller, but similar, effects between study arms (Figure 1B, Table 2). The AUC ratio of rfMDA was 0.83 (95%CI 0.72 – 0.94, p=0.002) relative to RACD, and for RAVC, the AUC ratio was 0.85 (95%CI 0.73 – 0.97, p=0.014) relative to non-RAVC. The AUC ratio for rfMDA plus RAVC was 0.70 (95% CI 0.58 - 0.85, p<0.001) relative to RACD only. A difference in antibody responses to *Pf*AMA1 between study arms was also observed based on unadjusted AUC values, with an AUC ratio of 0.73 (95%CI 0.48 – 0.97, p=0.028) for RAVC relative to non-RAVC, and an AUC ratio of 0.71 (95%CI 0.41 – 1.00, p=0.048) for rfMDA plus RAVC relative to RACD only. The distribution of cluster-level AUC values is shown by study arm in Supplementary Figure S5.

**Table 2.** Etramp5.Ag1 Area under the antibody acquisition curve (AUC) by intervention. Reference arms are clusters in the RACD, non-RAVC, or RACD only arms. Ratio of log AUC values in the intervention vs reference arms are estimated using generalised linear models (log link, gaussian family, and GEE for clustering at the EA-level) and adjusted for EA incidence in 2016, proportion of EA index cases covered, proportion of target population covered, median time to intervention, and distance from villages receiving an MOHSS intervention.

|  |  |  | Unadjusted |  | Adjusted |  |
| --- | --- | --- | --- | --- | --- | --- |
|  | Number of | Mean AUC value | AUC ratio | p-value | AUC ratio | p-value |
| **Human reservoir** |  |  |  |  |  |  |
| RACD (reference)* | 27 | 34,595 (31,496 – 37,693) | 1.00 | -- | 1.00 | -- |
| rfMDA* | 28 | 28,337 (25,429 – 31,244) | 0.82 (0.71 – 0.93) | 0.002 | 0.83 (0.72 – 0.94) | 0.002 |
| **Mosquito reservoir** |  |  |  |  |  |  |
| No RAVC (reference)† | 27 | 33,211 (30,074 – 36,348) | 1.00 | -- | 1.00 | -- |
| RAVC† | 28 | 29,382 (26,516 – 32,248) | 0.88 (0.76 – 1.00) | 0.060 | 0.85 (0.73 – 0.97) | 0.014 |
| **Human and mosquito reservoir** |  |  |  |  |  |  |
| RACD only (reference) | 13 | 35,449 (31,307 – 40,138) | 1.00 | -- | 1.00 | -- |
| rfMDA plus RAVC | 14 | 25,553 (21,060 – 31,005) | 0.72 (0.59 – 0.87) | 0.001 | 0.70 (0.58 – 0.85) | <0.001 |

\* With or without RAVC † With either RACD or rfMDA

### Serology-based coefficient of variation and sample size

The estimated sero-prevalence coefficient of variation between clusters, k, in RACD only clusters was 0.24, compared to a qPCR prevalence coefficient of variation of 0.71 (Figure 3A and 3B). For a range of cluster sizes, the number of clusters per arm was estimated to detect an effect size of either a 75% or a 50% reduction in sero-prevalence with 80% study power and a 5% significance level (alpha=5%), assuming a baseline sero-prevalence of 0.28 (mean sero-prevalence RACD only arm, Table 1). Assuming an average cluster sample of 65 individuals (based on the mean number of respondents per cluster in the cross-sectional survey), the minimum sample size required to observe a 75% reduction in sero-prevalence (between rfMDA plus RAVC vs RACD only) was 2.6 clusters per arm, while 5.4 clusters per arm would be required to observe a 50% reduction in sero-prevalence (between either rfMDA vs RACD or RAVC vs. no RAVC). The observed reductions in sero-prevalence to Etramp5.Ag1 from the study were 35% in the rfMDA plus RAVC compared to RACD only arms and 25% in the rfMDA compared to RACD arms, which would require a serology sample size of 11 and 22 clusters, respectively.

**Figure 3.**
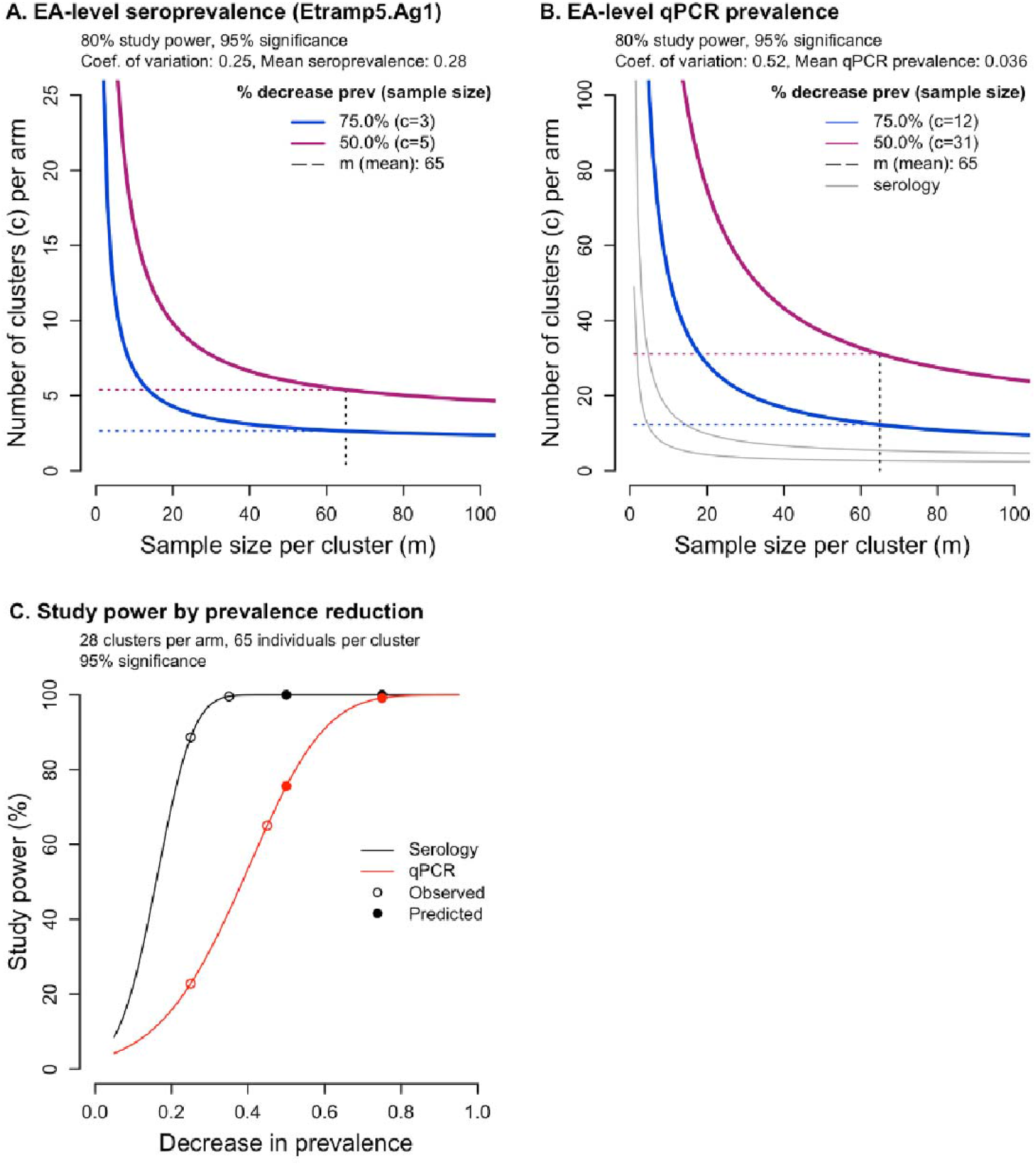
Coefficient of variation, k, and number of clusters per arm, c, using serology compared to qPCR as a trial endpoint. Number of clusters per arm is estimated for serology (A) and qPCR (B) based on predicted decrease in prevalence of 75% for rfMDA plus RAVC arm (blue) and 50% in rfMDA or RAVC arms (magenta). Mean cluster sample size, m (mean), is indicated by the dotted vertical black line, and the associated number of clusters required indicated by the horizontal dotted lines. Change in study power by decrease in prevalence (C) is shown for serology (black) and qPCR (red), with study power for predicted and observed decreases in prevalence indicated by filled and empty circles.

Alternatively, for the trial as designed with 28 clusters per arm to compare the two main effects (rfMDA vs RACD, and RAVC vs no RAVC), and 14 clusters per arm to compare the combination rfMDA plus RAVC vs RACD only, there would have been >99.9% power to detect reductions in seroprevalence of 50% and 75% respectively for each of the comparisons (i.e., virtually no chance of a type 2 error). Observed reductions in sero-prevalence of 35% and 25% can be detected with study power of 99.6% and 88.7% respectively (Figure 3C).

By contrast, assuming a baseline qPCR prevalence of 0.033 (mean prevalence in RACD only clusters), a minimum of 12.9 and 33 clusters per arm would be required to detect a decrease of 75% and 50% qPCR prevalence, respectively. Based on modelled relationship between log odds of sero-positivity and log odds of qPCR positivity (Supplementary Materials equation 1), a qPCR prevalence of 0.033 translates to Etramp5.Ag1 sero-prevalence of 0.25 (95% CrI 0.23 – 0.28), aligned with the observed baseline sero-prevalence in the RACD only arms noted above. In the study, the qPCR prevalence reductions observed were 45% in the rfMDA plus RAVC arms and 25% in the rfMDA arms.

Sensitivity analysis of the effect of coefficient of variation and baseline prevalence on required sample size was explored for both serology and qPCR endpoints (Figure 4A and 4B). For serology, based on coefficient of variation values between 0.1 to 0.5 and baseline sero-prevalence from 0.05 to 0.45, the estimated sample size was between 2.4 and 24.7 clusters per arm. For qPCR, based on coefficient of variation values between 0.5 to 0.9 and baseline qPCR prevalence from 0.02 to 0.10, estimated sample sizes ranged from 17.4 to 68.3 clusters per arm. When using 28 clusters per study arm, while there is 98% study power to detect a 75% reduction in qPCR prevalence, this study power drops to 72.9% when trying to detect a 50% reduction (Figure 3C). For the reductions in qPCR prevalence observed in the end-line survey, there is a 62.3% and 21.7% study power to detect a 35% and 25% decrease in prevalence, respectively.

**Figure 4.**
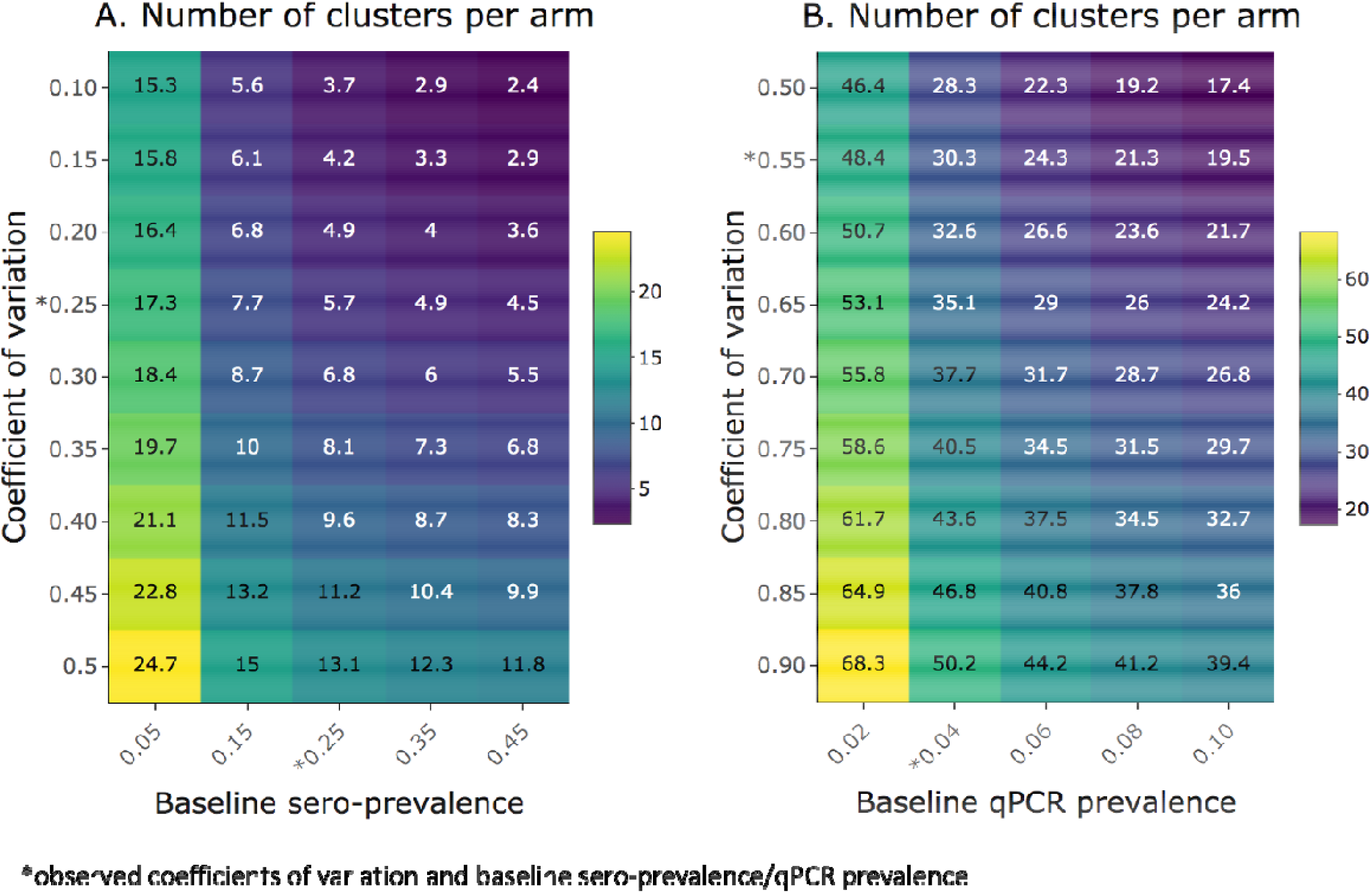
Number of clusters per arm, c, for a range of baseline prevalence and coefficient of variation values. Heatmaps show the number of clusters per arm required for a range of coefficient of variation values and sero-prevalence (A) or qPCR prevalence (B), assuming an average of 65 individuals per cluster and 50% reduction in sero- or qPCR-prevalence. Observed coefficients of variation and baseline sero- and qPCR-prevalence are indicated by asterisks.

## Discussion

This study sought to assess the application of serological markers for malaria exposure in the context of a cluster randomised trial, which was originally analysed using clinical incidence and qPCR prevalence as outcomes. Previous analysis showed that the introduction of rfMDA and RAVC, independently and in combination, had a significant effect on malaria clinical incidence and qPCR prevalence, after adjusting for factors that could not be balanced between study arms (as was done in Hsiang et al). This observation was replicated when assessing the efficacy of the interventions using the serological marker Etramp5.Ag1; rfMDA and RAVC were shown to be associated with significantly reduced antibody responses as independent and combined interventions. The intervention effects on antibody responses were detected using both binary and continuous antibody measurements.

The use of sero-incidence markers to evaluate trial outcomes was found to be comparable to the use of qPCR parasite prevalence, with the added benefit of increased precision in the measure of effect size (a standard error for sero-prevalence ratio of only 0.06 compared to a standard error of 0.19 for qPCR prevalence ratio). This is likely due to lower within-season fluctuations in population antibody responses compared to variations in levels of parasitaemia affecting detection by molecular assays. In previous studies in The Gambia, Etramp5.Ag1 antibody responses was found to persist for several months following the transmission season, before waning prior to transmission season.^23^ This suggests that serology is a more temporally stable measurement of malaria exposure throughout the transmission season but is still able to detect short-term changes occurring over the course of less than a year. This improved measurement consistency is reflected in a between-cluster coefficient of variation for sero-prevalence that is two-thirds lower than the coefficient of variation for qPCR prevalence. This can translate to significantly improved study power to detect fine-scale changes in transmission with smaller sample sizes. In multi-intervention studies, smaller effects are likely envisaged between study arms. For example, a number of trials are currently comparing the use of MDA with or without ivermectin,^32^ as well as the use of the RTS,S malaria vaccine combined with seasonal malaria chemoprevention (SMC).^33^ These studies will likely be evaluated in the context of standard malaria control intervention already in use and detecting subtle differences may be challenging.

Serological endpoints may be particularly relevant in the design of cluster randomised trials in low transmission settings with limitations in study power. In these areas, rates of clinical incidence or malaria infection prevalence may be very low or close to zero and would require prohibitively large sample sizes to detect intervention effects. This is highlighted in this study in Namibia, where all clusters measuring a qPCR prevalence or clinical incidence of zero still had detectable sero-positive individuals (Supplementary Figure S7). Serology may also serve as a cost-effective secondary endpoint to other measures of infection such as PCR.

Ongoing research to develop new serological markers into point-of-care lateral flow devices may enable a new method for rapid field-based malaria surveillance. This is increasingly important in settings such as Namibia to monitor for potential outbreaks and to prevent reintroduction of malaria infection following elimination. For Namibia, serological surveillance could be beneficial given the fluctuations in malaria incidence in recent years. Due to frequent population movement from neighbouring countries, especially along the Angolan and Zambian borders, low to moderate transmission has been found to persist and receptivity remains high.^4^

Our study findings have shown that the combination of higher prevalence and lower between cluster variation on sero-positivity compared to parasite positivity translate into substantially higher study power for serological endpoints, and near negligible chance of a type 2 error. Further research can help to operationally translate serology into a standardised trial tool. There have been only a limited number of studies measuring serology in malaria intervention trials, most of which have been in sub-Saharan Africa (Zambia^34^ and this study in Namibia). There have been studies in Haiti, Uganda, Zambia, The Gambia and Indonesia, where similar levels of immune responses to Etramp5.Ag1 have been measured, and further testing of this marker in the context of intervention trials could be useful.

One potential challenge is that rates of antibody acquisition, boost, and decay may differ between antigens and individuals. This may be one reason that strong antigenic signals were not observed for non-Etramp5.Ag1 markers in this study. Sequence variations in parasite proteins between populations or regions may also lead to some variation in antibody responses. Fortunately, refining the design of recombinant proteins for more precise serological surveillance is the subject of ongoing work.^35,36^ Improvements in assay design are currently leveraging multiplexing technology to measure the combined response to diverse panels of antigenic variants, capturing the full breadth of antibody responses in a population for single diseases or across multiple pathogens. Other strategies include the production of chimeric proteins with specific epitopes to these variants.

Biological variations in antibody responses are inevitable to some degree and integrating serological data with other important measures, such as age and transmission intensity, allows a comprehensive characterisation of epidemiological trends. For standardised surveillance across locations, panels based on studies in a diversity of settings will be beneficial. Serological surveillance is increasingly being standardised for a number infectious diseases such as dengue^37^, trachoma^38^, and lymphatic filariasis^39^. The inclusion of malaria in multi-disease sero-diagnostic panels^40,41^ can allow for cost and time efficiencies, reducing the number of single-disease surveys required in diverse health monitoring programmes. This can help guide integrated programme delivery rather than a reliance on multiple vertical programmes delivered separately.

Serological endpoints in intervention trials have become accepted as outcomes for other infectious diseases, such as arboviruses. For example, an ongoing cluster randomised trial on the efficacy of *Wolbachia*-infected *Aedes aegypti* in Brazil will measure reductions in sero-incidence to dengue, zika, and/or chikungunya virus as primary trial endpoints.^42^ In a similar randomised controlled trial in Indonesia, baseline sero-prevalence was used to infer age-specific transmission rates and median age to first infection to inform trial design.^43^ In the case of Zika, it has been suggested that the use of select antigens could help distinguish between antibody responses arising from vaccination versus natural infection, offering advantages over molecular diagnostics for trial evaluation and sampling.^44^ The findings from this study, together with ongoing innovations in assay design and multi-disease platforms, illustrate the potential application of serological markers as endpoints in randomised trials, especially in settings where measures of clinical incidence or infection may be less reliable due to limitations in health care seeking, or incomplete testing and reporting.

## Contributors

MSH, IK, and RG conceptualised and designed the study. MSKD, AB, JLS contributed to study design. MSH and RG provided overall oversight of the study. KWR and HN led the field implementation of the trial. LMP, CSG, and VS supported trial field coordination. LMP, LS, LW, and JY led the cross-sectional survey. PU and SK supported collaboration with the Namibia Ministry of Health and Social Services. KKAT and DM led the laboratory activities. CP, TH, and JG conducted the serological testing. MT and LMP conducted the molecular testing. LMP, BG, and CD provided additional oversight of the laboratory activities. LW led data management and analyses of serological data. BW supported data analyses. IK, MSH, and CD advised on the data analyses. LW wrote the manuscript. All authors contributed to data interpretation and approved the final draft of the manuscript.

## Data Availability

After publication, data collected from this study are available upon request to the corresponding author. Available data include de-identified individual participant data, cluster-level data, and a data dictionary defining each field in the set. A published manuscript of the protocol is also available online. Requests to conduct analyses outside the scope of this publication will be reviewed by the principal investigators (MSH, DM, RG, and IK) to determine whether a requester's proposed use of the data is scientifically and ethically appropriate and does not conflict with constraints or informed consent limitations identified by the institutions that granted ethical approval for the study. Requests to reanalyse the data presented in this Article will not require such review.

## Acknowledgements

The authors would like to thank the residents of Zambezi region who consented and participated in the study. We thank the field staff and the field supervisors (Miriam Sezuni, Sylvester Majakube, Simataa Nyati, Flavian Libita, and Tererai Msakwa). We thank Tanga Kashupi, Petrina Shikonde, and Munyaradzi Tambo, for supporting laboratory data collection and Patrick McCreesh for supporting data analysis.

## References

1. WHO | World malaria report 2017. WHO (2018).

2. Elimination 8. Annual Report 2016. (2016).

3. Smith Gueye, C. et al. Namibia’s path toward malaria elimination: a case study of malaria strategies and costs along the northern border. BMC Public Health 14, 1190 (2014).

4. Smith, J. L. et al. Malaria risk in young male travellers but local transmission persists: a case– control study in low transmission Namibia. Malar. J. 16, 70 (2017).

5. Noor, A. M. et al. Malaria Control and the Intensity of Plasmodium falciparum Transmission in Namibia 1969–1992. PLoS One 8, e63350 (2013).

6. Chanda, E. et al. Strengthening tactical planning and operational frameworks for vector control: the roadmap for malaria elimination in Namibia. Malar. J. 14, 302 (2015).

7. Chanda, E. et al. An investigation of the Plasmodium falciparum malaria epidemic in Kavango and Zambezi regions of Namibia in 2016. Trans. R. Soc. Trop. Med. Hyg. 112, 546–554 (2018).

8. WHO Global Malaria Programme. WHO malaria terminology. (2016).

9. Wu, L. et al. Comparison of diagnostics for the detection of asymptomatic Plasmodium falciparum infections to inform control and elimination strategies. Nature 528, S86–93 (2015).

10. Bousema, T., Okell, L., Felger, I. & Drakeley, C. Asymptomatic malaria infections: detectability, transmissibility and public health relevance. Nat. Rev. Microbiol. 12, 833–40 (2014).

11. McCreesh, P. et al. Subpatent malaria in a low transmission African setting: A cross-sectional study using rapid diagnostic testing (RDT) and loop-mediated isothermal amplification (LAMP) from Zambezi region, Namibia. Malar. J. 17, 480 (2018).

12. Cook, J. et al. Mass Screening and Treatment on the Basis of Results of a Plasmodium falciparum-Specific Rapid Diagnostic Test Did Not Reduce Malaria Incidence in Zanzibar. J. Infect. Dis. 211, 1476–1483 (2015).

13. Parker, D. M. et al. Limitations of malaria reactive case detection in an area of low and unstable transmission on the Myanmar–Thailand border. Malar. J. 15, 571 (2016).

14. Hsiang, M. S. et al. Active Case Finding for Malaria: A 3-Year National Evaluation of Optimal Approaches to Detect Infections and Hotspots Through Reactive Case Detection in the Low-transmission Setting of Eswatini. Clin. Infect. Dis. (2019). doi:10.1093/cid/ciz403

15. Hsiang, M. S. et al. Effectiveness of reactive focal mass drug administration and reactive focal vector control to reduce malaria transmission in the low malaria-endemic setting of Namibia: a cluster-randomised controlled, open-label, two-by-two factorial design trial. Lancet 395, 1361–1373 (2020).

16. Drakeley, C. J. et al. Estimating medium-and long-term trends in malaria transmission by using serological markers of malaria exposure. Proc. Natl. Acad. Sci. U. S. A. 102, 5108–13 (2005).

17. Cook, J. et al. Serological markers suggest heterogeneity of effectiveness of malaria control interventions on Bioko Island, equatorial Guinea. PLoS One 6, e25137 (2011).

18. Biggs, J. et al. Serology reveals heterogeneity of Plasmodium falciparum transmission in northeastern South Africa: implications for malaria elimination. Malar. J. 16, 48 (2017).

19. Wu, L. et al. Antibody responses to a suite of novel serological markers for malaria surveillance demonstrate strong correlation with clinical and parasitological infection across seasons and transmission settings in The Gambia. BMC Med. 18, 304 (2020).

20. Dewasurendra, R. L. et al. Effectiveness of a serological tool to predict malaria transmission intensity in an elimination setting. BMC Infect. Dis. 17, 49 (2017).

21. Drakeley, C. & Cook, J. Chapter 5. Potential contribution of sero-epidemiological analysis for monitoring malaria control and elimination: historical and current perspectives. Adv. Parasitol. 69, 299–352 (2009).

22. Helb, D. A. et al. Novel serologic biomarkers provide accurate estimates of recent Plasmodium falciparum exposure for individuals and communities. Proc. Natl. Acad. Sci. U. S. A. 112, E4438–47 (2015).

23. Wu, L. et al. Sero-epidemiological evaluation of malaria transmission in The Gambia before and after mass drug administration. BMC Med. 18, 331 (2020).

24. University of California, S. F. Evaluation of Targeted Parasite Elimination (TPE) in Namibiae. ClinicalTrials.cov Available at: https://clinicaltrials.gov/ct2/show/NCT02610400.

25. Medzihradsky, O. F. et al. Study protocol for a cluster randomised controlled factorial design trial to assess the effectiveness and feasibility of reactive focal mass drug administration and vector control to reduce malaria transmission in the low endemic setting of Namibia. BMJ Open 8, e019294 (2018).

26. National malaria case management guidelines. (Republic of Namibia, Ministry of Health and Social Services, 2014).

27. Wu, L. et al. Optimisation and standardisation of a multiplex immunoassay of diverse Plasmodium falciparum antigens to assess changes in malaria transmission using sero-epidemiology. Wellcome Open Res. 4, 26 (2019).

28. Nibsc. First WHO Reference Reagent for Anti-malaria (Plasmodium falciparum) human serum.

29. Ondigo, B. N. et al. Estimation of Recent and Long-Term Malaria Transmission in a Population by Antibody Testing to Multiple Plasmodium falciparum Antigens. J. Infect. Dis. 210, 1123– 1132 (2014).

30. Drakeley, C. J. et al. Estimating medium- and long-term trends in malaria transmission by using serological markers of malaria exposure. Proc. Natl. Acad. Sci. U. S. A. 102, 5108–13 (2005).

31. Hayes, R. & Moulton, L. Cluster Randomised Trials. (Chapman & Hall/CRC Press, 2009).

32. Dabira, E. D. et al. Mass drug administration with high-dose ivermectin and dihydroartemisinin-piperaquine for malaria elimination in an area of low transmission with high coverage of malaria control interventions: Protocol for the massiv cluster randomized clinical trial. JMIR Res. Protoc. 9, (2020).

33. Chandramohan, D. et al. Seasonal malaria vaccination: protocol of a phase 3 trial of seasonal vaccination with the RTS,S/AS01E vaccine, seasonal malaria chemoprevention and the combination of vaccination and chemoprevention. BMJ Open 10, e035433 (2020).

34. Bridges, D. J. et al. Community-led Responses for Elimination (CoRE): A study protocol for a community randomized controlled trial assessing the effectiveness of community-level, reactive focal drug administration for reducing Plasmodium falciparum infection prevalence and incidence in Southern Province, Zambia. Trials 18, (2017).

35. Longley, R. J. et al. Development and validation of serological markers for detecting recent Plasmodium vivax infection. Nat. Med. 26, 741–749 (2020).

36. Yman, V. et al. Distinct kinetics in antibody responses to 111 Plasmodium falciparum antigens identifies novel serological markers of recent malaria exposure. medRxiv (2020).

37. Immunization, Vaccines and Biologicals A Guide to the Design and Conduct of Dengue Serosurveys. (2017).

38. Martin, D. L. et al. The use of serology for trachoma surveillance: Current status and priorities for future investigation. PLoS Negl. Trop. Dis. 14, e0008316 (2020).

39. Won, K. Y. et al. Use of antibody tools to provide serologic evidence of elimination of lymphatic filariasis in the Gambia. Am. J. Trop. Med. Hyg. 98, 15–20 (2018).

40. Arnold, B. F. et al. Measuring changes in transmission of neglected tropical diseases, malaria, and enteric pathogens from quantitative antibody levels. PLoS Negl. Trop. Dis. 11, e0005616 (2017).

41. Arnold, B. F., Scobie, H. M., Priest, J. W. & Lammie, P. J. Integrated serologic surveillance of population immunity and disease transmission. Emerg. Infect. Dis. 24, 1188–1194 (2018).

42. A Cluster-randomized Trial to EValuate the Efficacy of Wolbachia-InfecTed Aedes Aegypti Mosquitoes in Reducing the Incidence of Arboviral Infection in Brazil (EVITA Dengue) - Full Text View - ClinicalTrials.gov. Available at: https://clinicaltrials.gov/ct2/show/NCT04514107. (Accessed: 22nd February 2021)

43. Indriani, C. et al. Baseline characterization of dengue epidemiology in Yogyakarta City, Indonesia, before a randomized controlled trial of wolbachia for arboviral disease control. Am. J. Trop. Med. Hyg. 99, 1299–1307 (2018).

44. Collins, M. H. Serologic tools and strategies to support intervention trials to combat Zika virus infection and disease. Tropical Medicine and Infectious Disease 4, (2019).

