## Supplementary Materials for "Serological evaluation of a cluster randomised trial on the use of reactive focal mass drug administration and reactive vector control to reduce malaria transmission in Zambezi Region, Namibia"

Supplementary Material

**Figure S1.** **Number of clusters by study arm**

|  |  | rfMDA vs. RACD | |
| --- | --- | --- | --- |
|  |  | **RACD**  **(28 clusters)** | **rfMDA**  **(28 clusters)** |
| RAVC vs  No RAVC | **No RAVC**  **(28 clusters)** | RACD only  (14 clusters) | rfMDA only  (14 clusters) |
|  | **RAVC**  **(28 clusters)** | RACD + RAVC  (14 clusters) | rfMDA + RAVC  (14 clusters) |

**Table S1. Demographics of study population by study arm**

|  | RACD only | RACD plus RAVC | rfMDA only | rfMDA plus RAVC |
| --- | --- | --- | --- | --- |
| Individuals | n=990 (%) | n=1003 (%) | n=915 (%) | n=749 (%) |
| Age group |  |  |  |  |
| 1 – 5 years | 182 (18.4) | 175 (17.5) | 143 (15.6) | 139 (18.5) |
| 6 – 15 years | 272 (27.5) | 277 (27.6) | 264 (28.9) | 214 (28.6) |
| >15 years | 536 (54.1) | 551 (54.9) | 508 (55.5) | 396 (52.9) |
| Fever ($\boldsymbol{\geq}$37.5°C) | 4 (0.4) | 6 (0.6) | 7 (0.8) | 6 (0.8) |
| Sex |  |  |  |  |
| Female | 555 (56.1) | 562 (56.0) | 499 (54.5) | 400 (53.4) |
| Male | 435 (43.9) | 441 (44.0) | 416 (45.5) | 349 (46.6) |
| Slept outdoors in past 2 weeks | 52 (5.3) | 51 (5.1) | 64 (7.0) | 48 (6.4) |
| Slept under bed net previous night | 220 (22.2) | 195 (19.5) | 168 (18.4) | 198 (26.5) |
| Household reported IRS in past 12 months | 811 (82.1) | 840 (85.8) | 617 (68.6) | 538 (73.9) |
| Travel in past 8 weeks | 104 (10.5) | 117 (11.7) | 118 (12.9) | 122 (16.3) |

Table S2. Summary of antigen constructs and coupling conditions in multiplex Luminex panel.

| Gene ID | Antigen | Strain | Antigen bead coupling concentration (ug/mL) | Purification Tag | Location | Description | Reference |
| --- | --- | --- | --- | --- | --- | --- | --- |
| PF3D7_0930300 | *Pf*MSP1_19_ | Wellcome | 42.31 | GST | Merozoite surface | 19kDa fragment of MSP1 molecule | ^1^ |
| PF3D7_1133400 | *Pf*AMA1 | FVO | 3.90 | His_x6_ | Sporozoite / Merozoite | Apical membrane antigen 1 | ^2^ |
| PF3D7_1035300 | *Pf*GLURP.R2 | F32 | 9.22 | N/A | Merozoite | Glutamate rich protein R2 | ^3^ |
| PF3D7_0731500 | EBA175 | 3D7 | 408.32 | GST | Merozoite | Erythrocyte binding antigen-175 | ^4^ |
| PF3D7_1335400 | Rh2.2030 | D10 | 244.30 | GST | Merozoite | Reticulocyte binding protein homologue 2 | ^5^ |
| PF3D7_0532100 | Etramp5.Ag1 | 3D7 | 34.93 | GST | iRBC/PVM | Early transcribed membrane protein 5 | ^6^ |
| PF3D7_0402400 | GEXP18 | 3D7 | 625.00 | GST | Gametocytes | Gametocyte exported protein 18 | ^7^ |
| PF3D7_0501100.1 | HSP40.Ag1 | 3D7 | 42.54 | GST | iRBC / Gametocytes | Heat shock protein 40, type II | ^7^ |
| -- | GST | -- | 85.99 | -- | -- | GST expression tag |  |
| -- | TT | -- | 61.52 | -- | -- | Tetanus Toxoid |  |

***infected red blood cell – iRBC; parasitophorous vacuole membrane – PVM, glutathione S –transferase - GST

Table S3. Antigen classification based on magnitude of association of sero-positivity with age. Individual-level odds of sero-positivity by age category is estimated using generalised linear models (log link, binomial family, GEE with clustering at EA-level), adjusted for age category (1-5 years, 5-15 years, >15 years), gender, fever, EA incidence in 2016, proportion of EA cases covered, median time to intervention, and distance from villages receiving an MOHSS intervention.

| Antigen | Adjusted odds ratio of seropositivity  by age (95%CI)* | | Sero-marker classification |
| --- | --- | --- | --- |
|  | **Ages 5-15 years** | **Ages >15 years** |  |
| Etramp5.Ag1 | 1.59 (1.29 – 1.95) | 1.80 (1.51 – 2.15) | Short-term marker |
| GEXP18 | 1.08 (0.92 – 1.26) | 1.30 (1.11 – 1.51) | Short-term marker |
| HSP40.Ag1 | 1.38 (1.08 – 1.78) | 2.74 (2.15 – 3.50) | Short-term marker |
| EBA175 | 1.69 (1.28 – 2.23) | 4.46 (3.52 – 5.64) | Medium-term marker |
| Rh2.2030 | 3.75 (2.52 – 5.58) | 14.55 (9.92 – 21.33) | Long-term marker |
| *Pf*MSP1_19_ | 3.10 (2.31 – 4.16) | 6.38 (4.67 – 8.71) | Long-term marker |
| *Pf*AMA1 | 2.57 (1.90 – 3.49) | 8.94 (6.62 – 12.06) | Long-term marker |
| *Pf*GLURP.R2 | 2.71 (1.73 – 4.23) | 7.43 (5.02 – 11.00) | Long-term marker |

*compared to reference age category of 1-5 years

Figure S2. Sero-prevalence by intervention and antigen. Unadjusted mean sero-prevalence by study arm and intervention, estimated using generalised linear models (log link, binomial family, GEE with clustering at EA-level). Mean sero-prevalence is shown for rfMDA vs RACD (black), RAVC vs. no RAVC (blue) and rfMDA plus RAVC vs. RACD only (magenta)

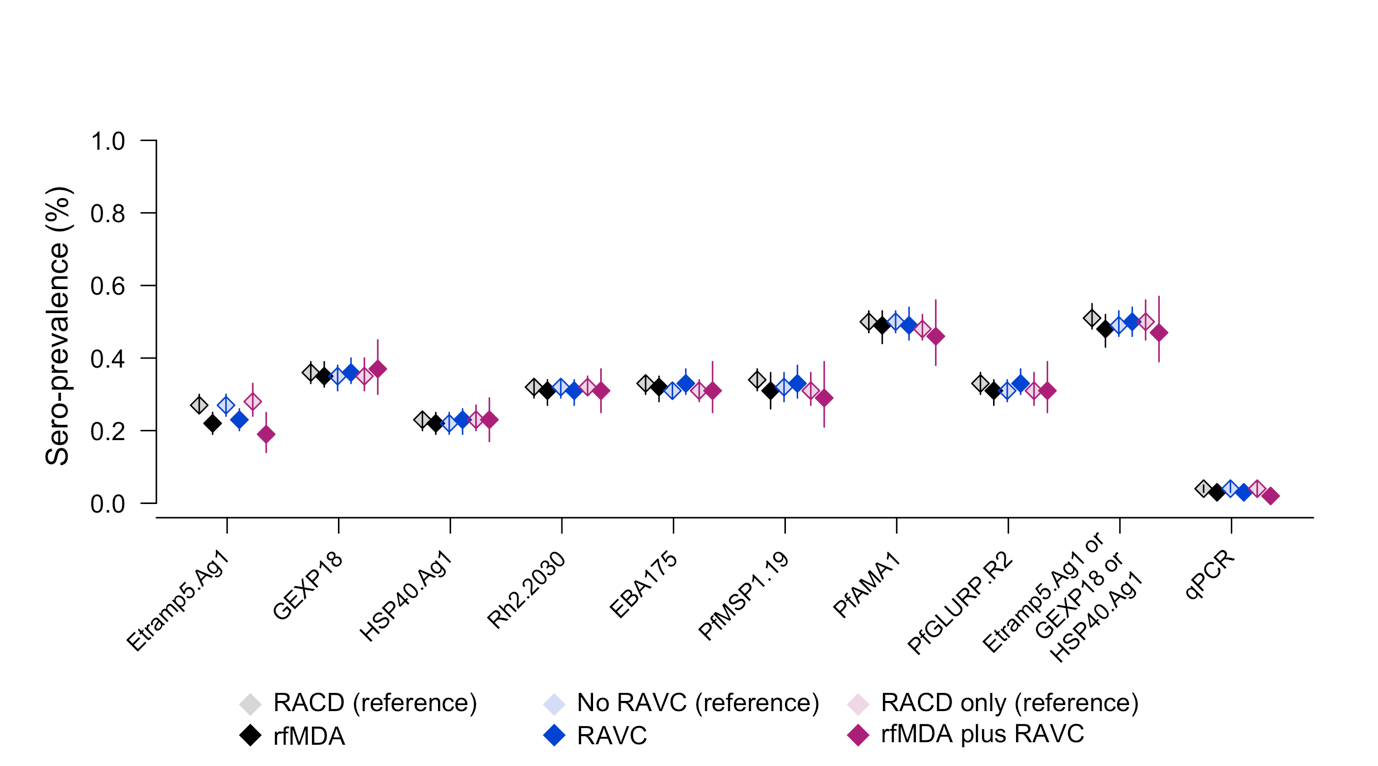

Table S4. Sero-prevalence by intervention and antigen. Mean sero-prevalence by antigen and intervention, estimated with generalised linear models (log link, binomial family, GEE with clustering at EA-level).

|  | Mean sero-prevalence (95%CI) | | | | |
| --- | --- | --- | --- | --- | --- |
|  | **Etramp5.Ag1** | **GEXP18** | **HSP40.Ag1** | **Etramp5.Ag1 or GEXP18 or HSP40.Ag1** | **qPCR** |
| RACD | 0.27 (0.25 - 0.30) | 0.36 (0.33 - 0.39) | 0.23 (0.20 - 0.25) | 0.51 (0.48 - 0.55) | 0.04 (0.03 - 0.05) |
| rfMDA | 0.22 (0.19 - 0.25) | 0.35 (0.31 - 0.39) | 0.22 (0.19 - 0.25) | 0.47 (0.43 - 0.52) | 0.03 (0.02 - 0.05) |
| No RAVC | 0.27 (0.24 - 0.30) | 0.35 (0.31 - 0.38) | 0.22 (0.19 - 0.25) | 0.49 (0.46 - 0.53) | 0.04 (0.03 - 0.06) |
| RAVC | 0.23 (0.20 - 0.26) | 0.36 (0.33 - 0.40) | 0.23 (0.19 - 0.26) | 0.50 (0.46 - 0.54) | 0.03 (0.02 - 0.04) |
| RACD only | 0.28 (0.24 - 0.33) | 0.35 (0.31 - 0.40) | 0.23 (0.20 - 0.27) | 0.50 (0.45 - 0.56) | 0.04 (0.02 - 0.06) |
| rfMDA plus RAVC | 0.18 (0.14 - 0.25) | 0.37 (0.29 - 0.45) | 0.23 (0.17 - 0.29) | 0.47 (0.39 - 0.56) | 0.02 (0.01 - 0.03) |
|  | **Rh2.2030** | **EBA175** | ***Pf*MSP1_19_** | ***Pf*AMA1** | ***Pf*GLURP.R2** |
| RACD | 0.31 (0.29 - 0.34) | 0.33 (0.30 - 0.35) | 0.34 (0.31 - 0.37) | 0.50 (0.47 - 0.53) | 0.33 (0.30 - 0.36) |
| rfMDA | 0.31 (0.27 - 0.34) | 0.32 (0.28 - 0.35) | 0.31 (0.26 - 0.36) | 0.49 (0.45 - 0.53) | 0.31 (0.27 - 0.34) |
| No RAVC | 0.32 (0.29 - 0.34) | 0.31 (0.29 - 0.33) | 0.32 (0.28 - 0.36) | 0.50 (0.47 - 0.53) | 0.31 (0.28 - 0.33) |
| RAVC | 0.31 (0.25 - 0.37) | 0.33 (0.30 - 0.37) | 0.33 (0.29 - 0.38) | 0.49 (0.45 - 0.53) | 0.33 (0.30 - 0.37) |
| RACD only | 0.32 (0.30 - 0.35) | 0.31 (0.28 - 0.34) | 0.31 (0.27 - 0.36) | 0.48 (0.45 - 0.52) | 0.31 (0.27 - 0.35) |
| rfMDA plus RAVC | 0.31 (0.25 - 0.37) | 0.31 (0.25 - 0.39) | 0.29 (0.21 - 0.38) | 0.46 (0.38 - 0.56) | 0.31 (0.25 - 0.39) |

Table S5. Etramp5.Ag1 sero-prevalence ratio by study arm and intervention. Sero-prevalence ratio by study arm and intervention, estimated using generalised linear models (log link, binomial family, GEE with clustering at EA-level).

|  | Unadjusted | | | Adjusted | | |
| --- | --- | --- | --- | --- | --- | --- |
|  | **PR** | **95% CI** | **p-value** | **PR** | **95% CI** | **p-value** |
| By study arm |  |  |  |  |  |  |
| RACD only (reference) | 1.00 | 1.00 |  | 1.00 | 1.00 |  |
| RACD + RAVC | 0.79 | 0.64 – 0.93 | 0.004 | 0.78 | 0.65 – 0.91 | <0.001 |
| rfMDA | 0.85 | 0.70 – 1.00 | 0.057 | 0.79 | 0.67 – 0.92 | 0.001 |
| rfMDA + RAVC | 0.65 | 0.49 – 0.87 | 0.003 | 0.59 | 0.46 – 0.76 | <0.001 |
| EA incidence 2016 | -- | -- | -- | 1.00 | 1.00 – 1.00 | <0.001 |
| Median time to intervention | -- | -- | -- | 1.00 | 0.98 – 1.01 | 0.576 |
| Mean EA coverage of index cases | -- | -- | -- | 0.82 | 0.46 – 1.44 | 0.483 |
| Mean EA coverage of target population |  |  |  | 0.98 | 0.53 – 1.82 | 0.955 |
| <500m from MOHSS intervention | -- | -- | -- | 1.41 | 1.18 – 1.67 | <0.001 |

* With or without RAVC	† With either RACD or rfMDA

Figure S3. Unadjusted mean AUC value by study arm and intervention, estimated using generalised linear models (log link, gaussian family, GEE with clustering at EA-level). Mean AUC is shown for rfMDA vs RACD (black), RAVC vs. no RAVC (blue) and rfMDA plus RAVC vs. RACD only (magenta)

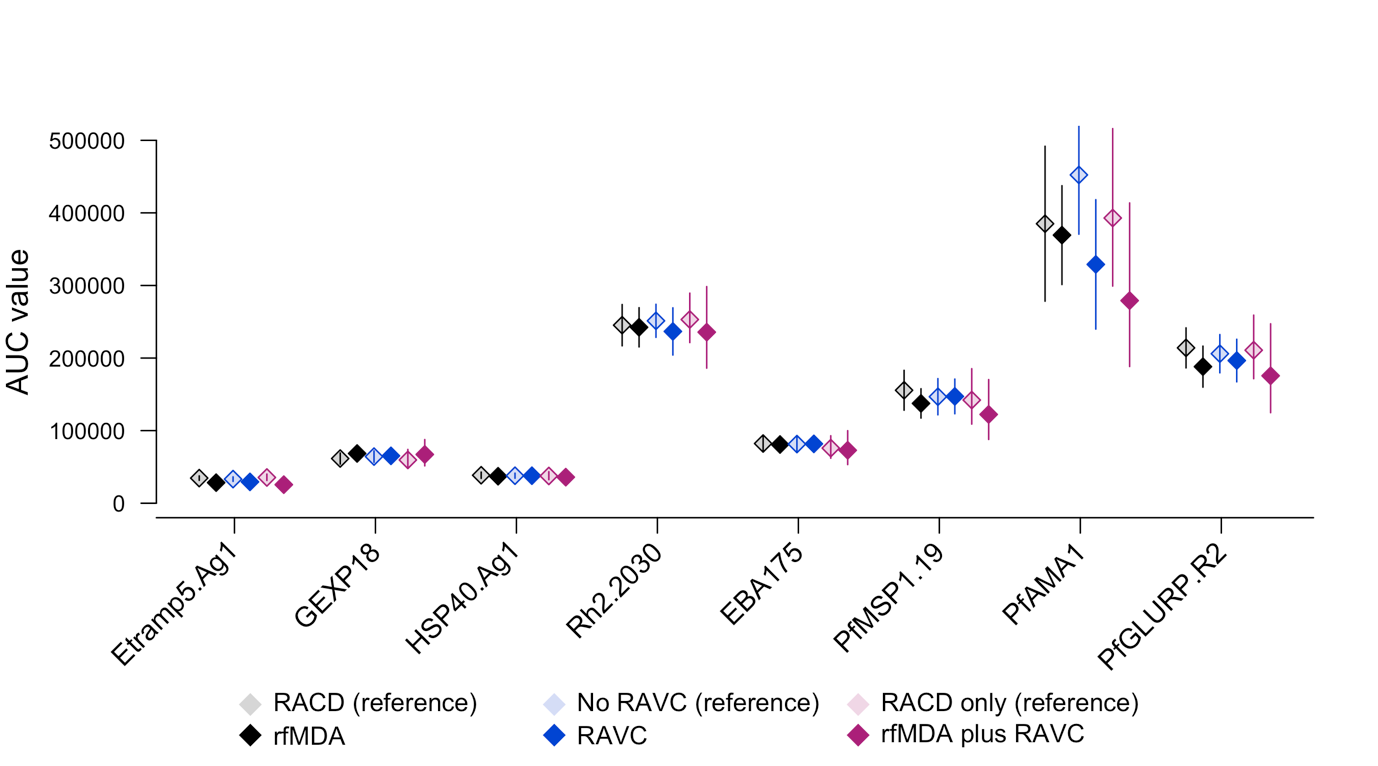

Table S6. Etramp5.Ag1 AUC ratio by study arm and intervention. Ratio of log AUC values by study are and intervention is estimated using generalised linear models (log link, gaussian family, inverse-weighted by 95%CI AUC) and adjusted for EA incidence in 2016, proportion of EA cases covered, median time to intervention, and distance from villages receiving an MOHSS intervention.

|  | Unadjusted | | | Adjusted | | |
| --- | --- | --- | --- | --- | --- | --- |
|  | **AUC ratio** | **95%CI** | **p-value** | **AUC ratio** | **95%CI** | **p-value** |
| By study arm |  |  |  |  |  |  |
| RACD only | 1.00 | 1.00 | -- | 1.00 | 1.00 | -- |
| RACD + RAVC | 0.81 | 0.71 – 0.93 | 0.002 | 0.82 | 0.72 – 0.94 | 0.002 |
| rfMDA only | 0.88 | 0.76 – 1.00 | 0.060 | 0.85 | 0.73 – 0.97 | 0.014 |
| rfMDA + RAVC | 0.72 | 0.59 – 0.87 | 0.001 | 0.70 | 0.58 – 0.85 | <0.001 |
| EA incidence 2016 | -- | -- | -- | 1.00 | 1.00 | <0.001 |
| Median time to intervention | -- | -- | -- | 1.00 | 0.99 – 1.01 | 0.843 |
| Proportion of EA cases covered |  |  |  | 0.93 | 0.60 – 1.46 | 0.762 |
| Mean enrolment proportion | -- | -- | -- | 1.20 | 0.63 – 2.27 | 0.574 |
| <500m from MOHSS intervention | -- | -- | -- | 1.16 | 0.98 – 1.38 | 0.091 |

Figure S4. Distribution of EA-level sero-prevalence for Etramp5.Ag1, by intervention. The distribution of cluster-specific sero-prevalence is compared in the rfMDA vs. RACD arms (black), RAVC vs. no RAVC arms (blue) and rfMDA plus RAVC vs. RACD only arms (magenta).

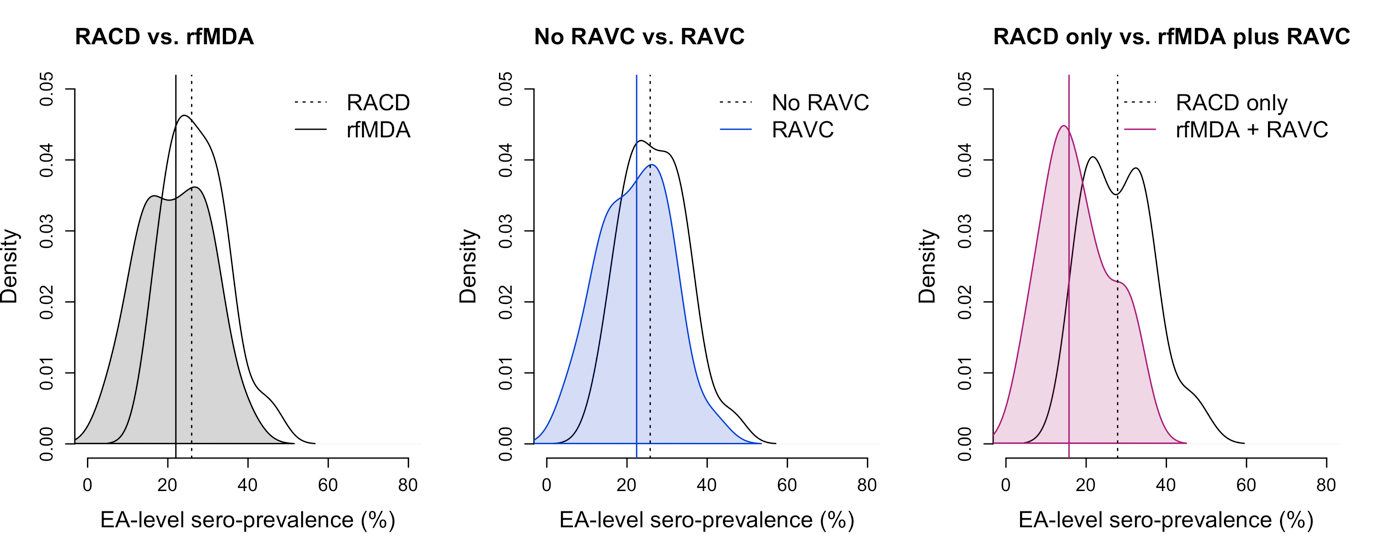

Figure S5. Distribution of EA-level AUCs for Etramp5.Ag1, by intervention. The distribution of cluster-specific AUC values is compared in the rfMDA vs. RACD arms (black), RAVC vs. no RAVC arms (blue) and rfMDA plus RAVC vs. RACD only arms (magenta).

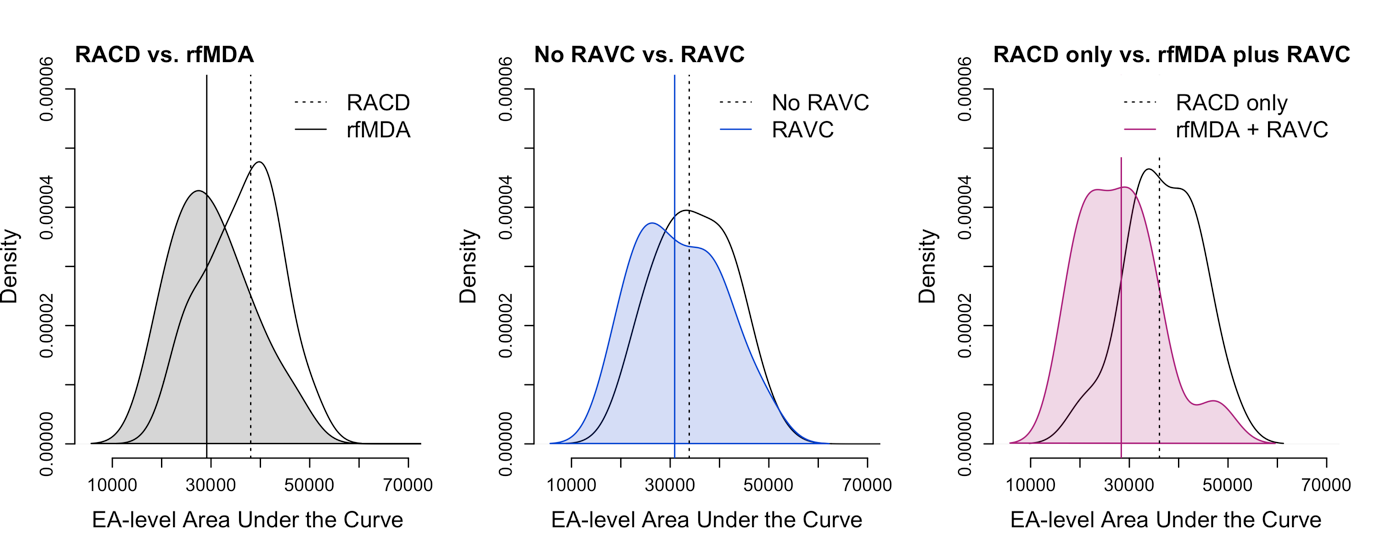

Figure S6. Etramp5.Ag1 antibody acquisition model fit by enumeration area.

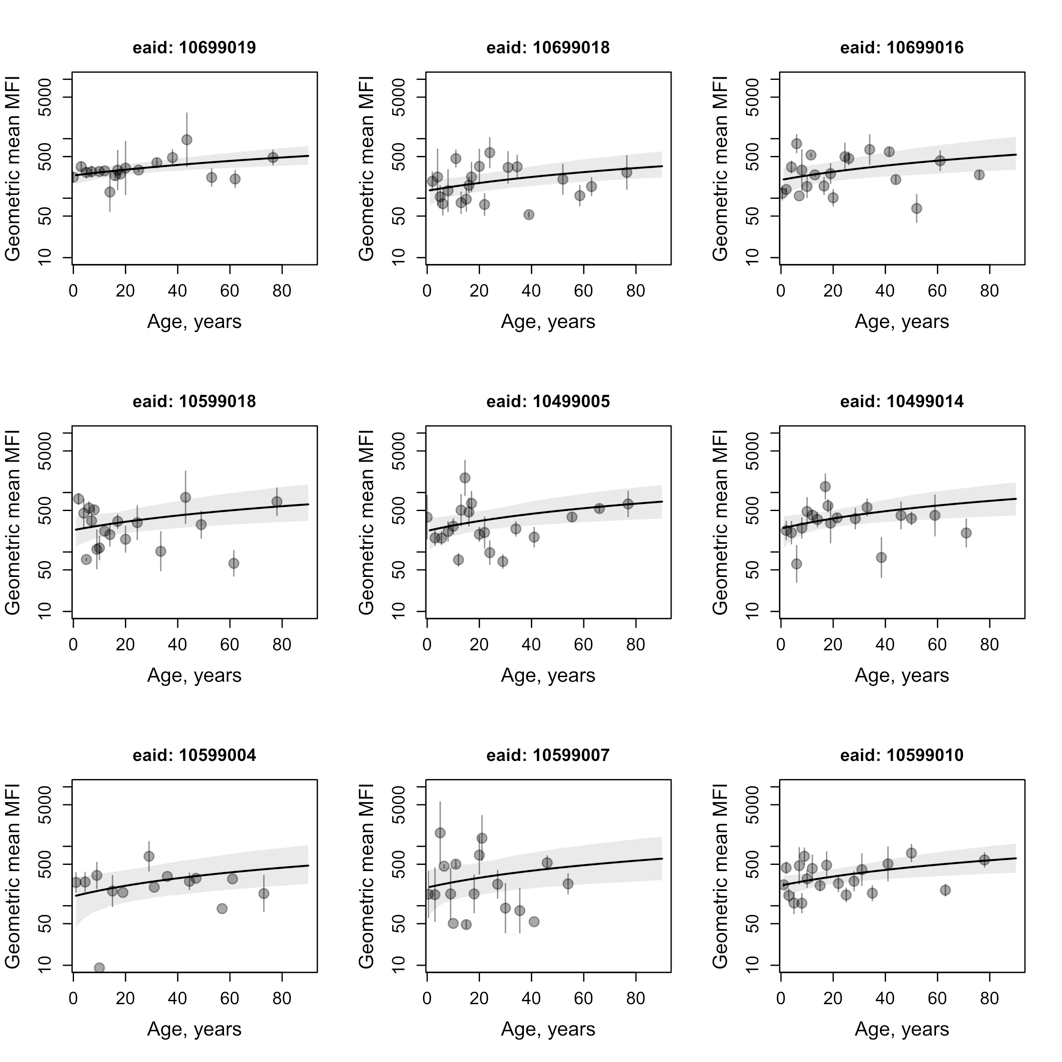

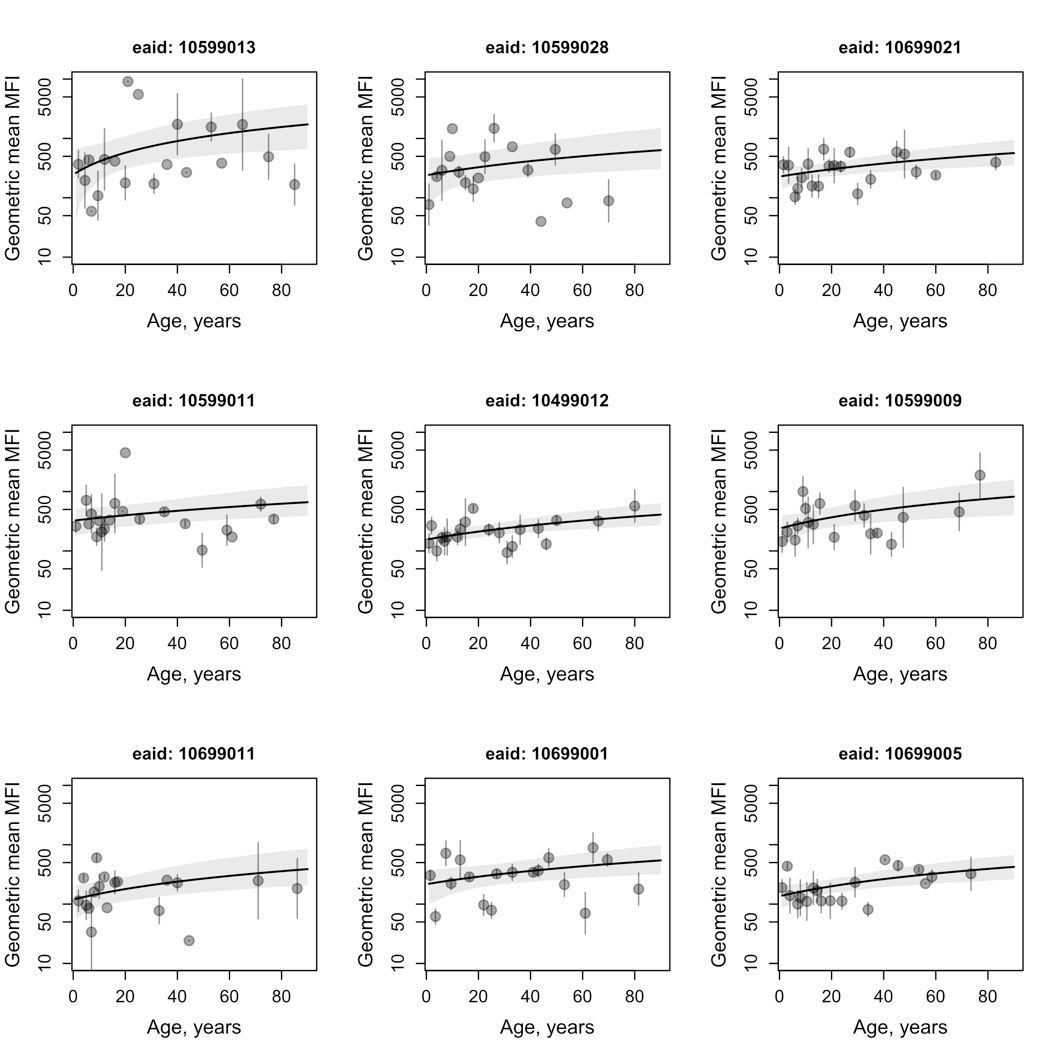

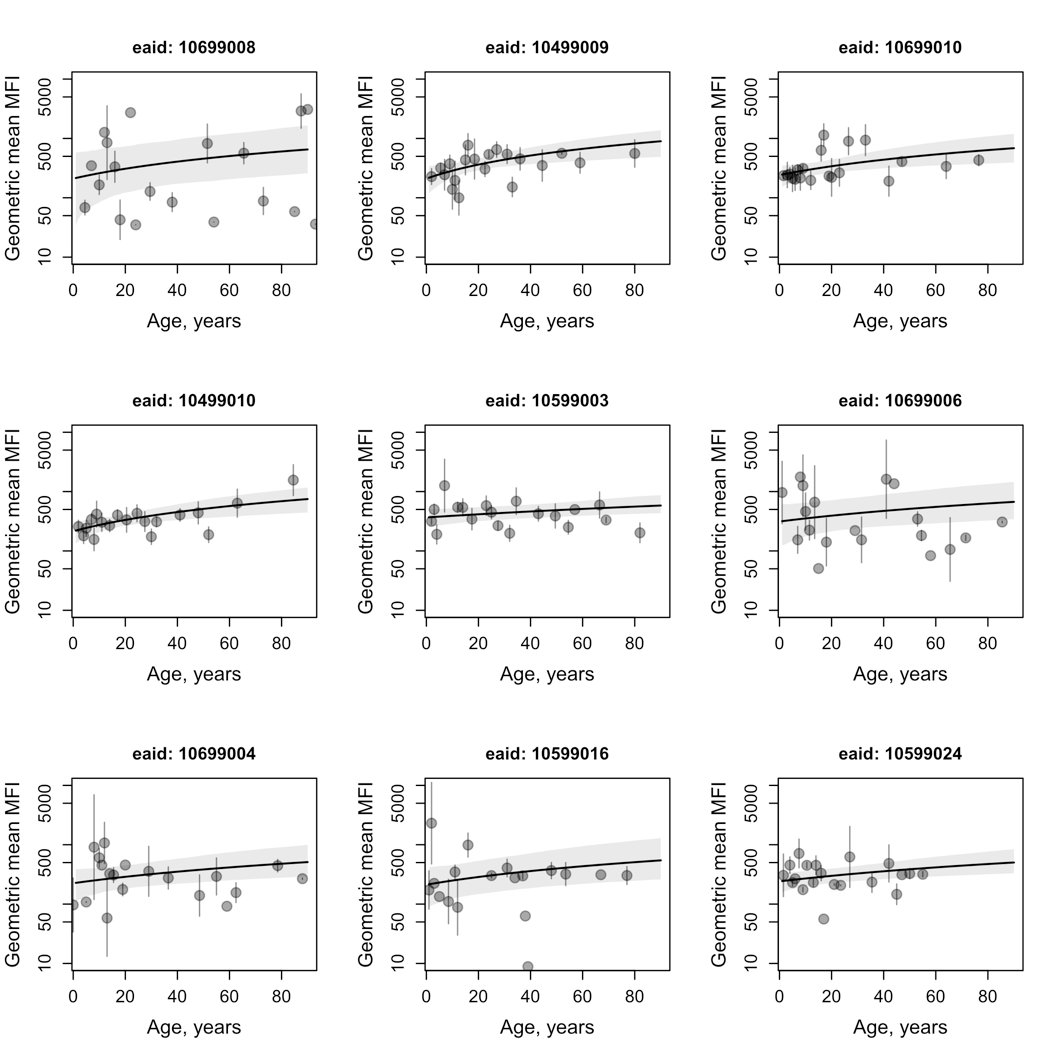

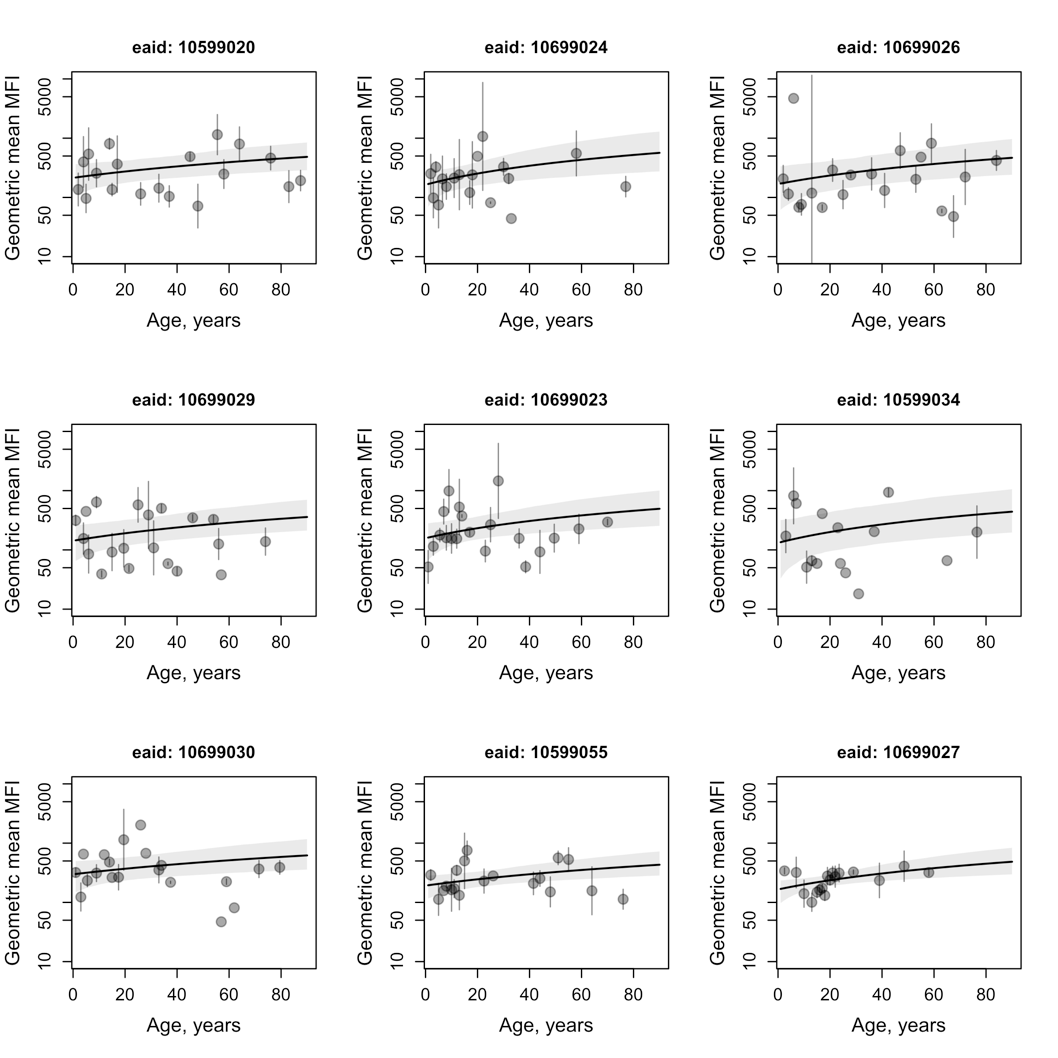

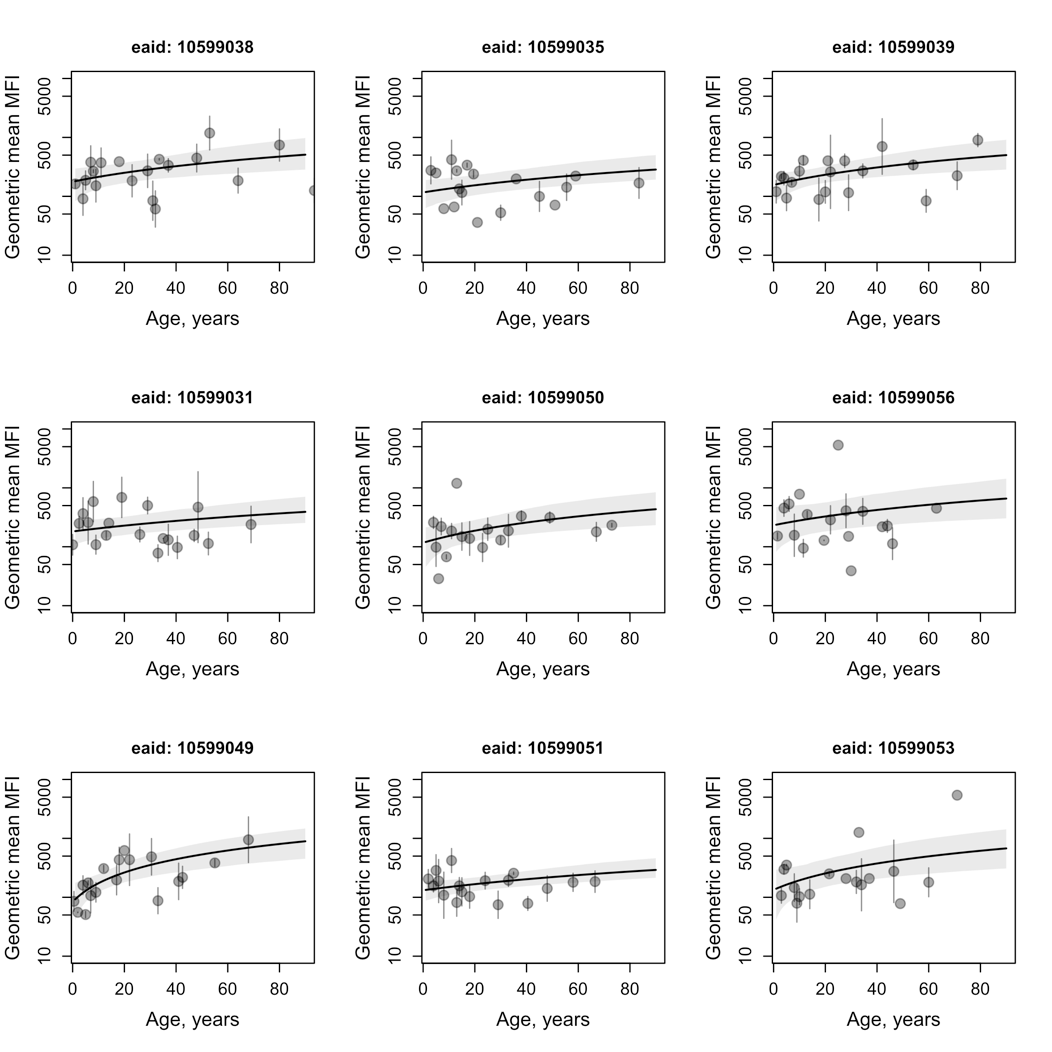

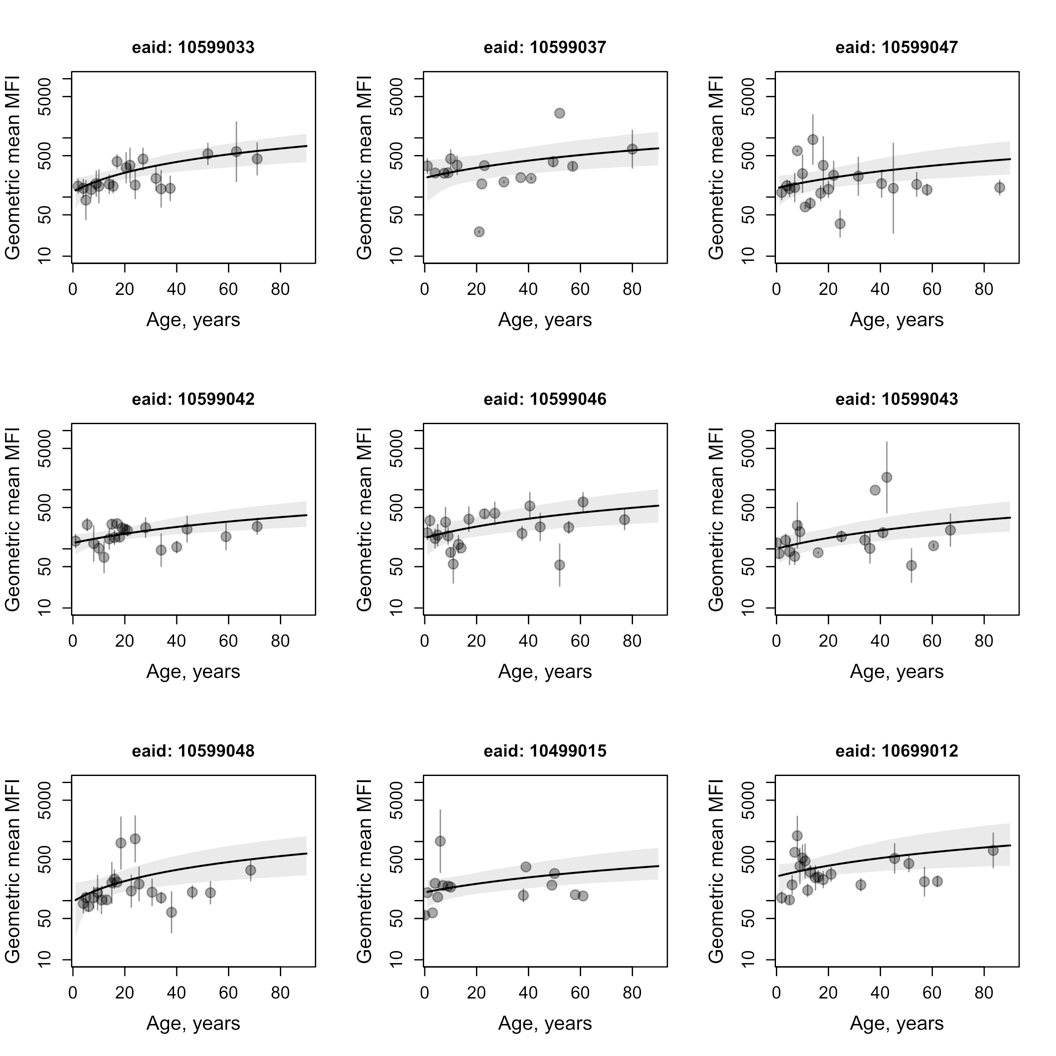

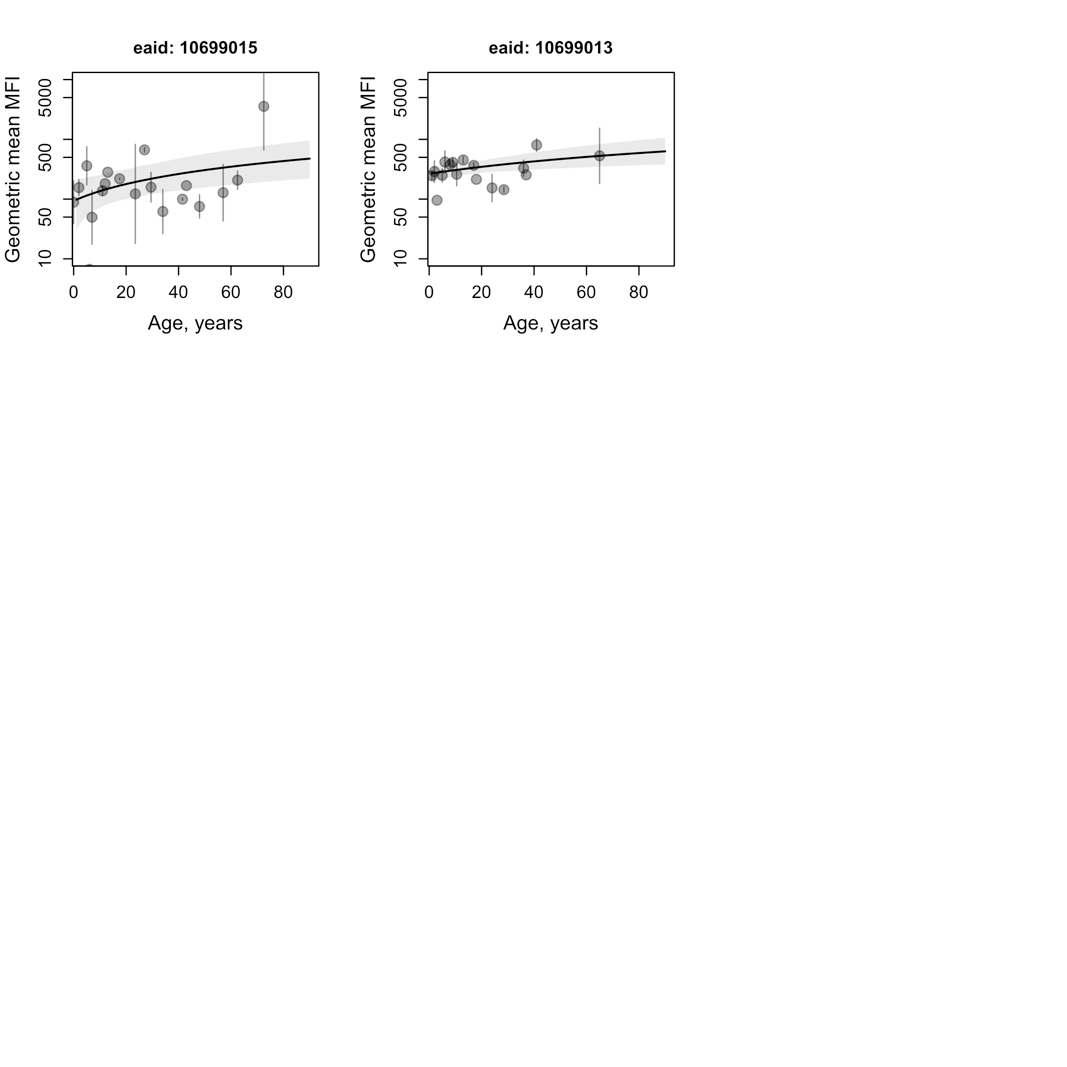

***Relationship between sero-prevalence, qPCR prevalence and clinical incidenc*e.**

At the individual level, the association between sero-positivity and concurrent qPCR positivity was estimated using a generalised linear model with GEE clustering at the EA-level and adjusted for age group, gender, fever, study arm, and EA clinical incidence in 2016.

Additionally, relationships between trial endpoints were estimated using cluster-level sero-prevalence, qPCR prevalence and clinical incidence. The association between sero-prevalence and qPCR prevalence was estimated by fitting a linear relationship on the log odds scale:

$\theta_{Si}= \alpha_{0}+ \beta_{0}{(\theta}_{Pi}- \bar{\theta}_{P})+ \beta_{1}\omega_{1}+ \beta_{2}\omega_{2}+ \beta_{3}\omega_{1}\omega_{2}$	(equation 1)

Where $\theta_{Si}$ is the log odds of sero-positivity measured in cluster i, $\theta_{Pi}$ is the log odds of qPCR positivity in cluster i, $\bar{\theta}_{P}$ is the mean log odds of qPCR prevalence across all clusters, $\omega_{1}$ and $\omega_{2}$ are binary variables equal to 1 for study arms receiving rfMDA and RAVC, respectively. $\alpha_{0}$ represents the expected log odds of sero-positivity when log odds of the PCR prevalence in cluster i is equal to the mean across all clusters, $\beta_{0}$ is the regression coefficient for the effect of PCR prevalence, $\beta_{1}$is the regression coefficient for the effect of rfMDA, $\beta_{2}$ is the regression coefficient for the effect of RAVC and $\beta_{3}$ is the regression coefficient for the interaction between rfMDA and RAVC.

To account for the influence of sample size and sampling variation across surveys, the probability of an individual being parasite-positive is binomially distributed based on the number of individuals parasite positive out of the total number of individuals tested in each cluster and defined as $\frac{e^{(\log odds)}}{1+e^{\log odds}}$ and log odds = $\log_{e} (\frac{prevalence}{1-prevalence})$.

Next, to estimate the relationship between sero-prevalence and clinical incidence, a linear relationship was also fitted between log sero-prevalence and log clinical incidence:

$\theta_{Si}= \alpha_{0}+ \beta_{0}{(\theta}_{Ci}- \bar{\theta}_{C}) + \beta_{1}\omega_{1}+ \beta_{2}\omega_{2}+ \beta_{3}\omega_{1}\omega_{2}$	(equation 2)

$\pi_{i} \sim Poisson(\frac{\varepsilon_{i}}{\rho_{i}})$	(equation 3)

Where $\theta_{Si}$ is the log sero-prevalence measured in cluster *i,* $\theta_{Ci}$ is the log of clinical incidence ($\pi_{i}$) in cluster *i*, $\bar{\theta}_{C}$ is the mean log of clinical incidence across all clusters, $\varepsilon_{i}$ is the number of clinical episodes in cluster *i,* and $\rho_{i}$ is the total person-years at risk (PYAR) in cluster *i*. $\alpha_{0}$is the expected log sero-prevalence when log of clinical incidence in cluster *i* is equal to the mean across all clusters, and $\beta_{0}$ is the regression coefficient for the effect of clinical incidence, and $\beta_{1}$, $\beta_{2}$, $\beta_{3}$ are the regression coefficients for the effect of rfMDA, RAVC and their interaction effect as described above. Cluster-specific clinical incidence is represented as the number of clinical episodes out of total person-years at risk and assumed to follow a Poisson distribution to account for between-cluster variation in PYAR.

Both models were fitted using Bayesian Markov Chain Monte Carlo (MCMC) methods in JAGS version 3.4.0 and the *rjags* package in R version 3.6.1.

***Relationship between trial endpoints sero-prevalence, qPCR prevalence and clinical incidence.*** At the individual-level, sero-positivity to Etramp5.Ag1 was significantly associated with an increase in concurrent qPCR positivity (aOR 1.61 95%CI 1.90 – 2.59, p=0.049) (Table S6). Based on the models described in equations 1-3, a quantitative association between three trial endpoints was established (Figure S6). Sero-prevalence was positively associated with both qPCR prevalence and clinical incidence. Cluster-level sero-prevalence ranged from 0.06 to 0.46, compared to a qPCR prevalence range of 0 to 0.45 and clinical incidence ranging from 0 to 20 events per 100 PYAR. Of the 17 clusters with qPCR prevalence of zero, sero-prevalence in the same clusters ranged from 0.06 to 0.35. For the 11 clusters measuring clinical incidence of zero, sero-prevalence ranged from 0.06 to 0.25. For both PCR prevalence and clinical incidence, the relationship with sero-prevalence was not significantly different by study arm. Mean posterior estimates for model parameters are included in Supplementary Table S8.

**Table S7. Individual-level association between qPCR sero-positivity and concurrent Etramp5.Ag1 sero-positivity (response variable).** Odds ratios are based on generalised linear model with GEE clustering at the EA-level and adjusted for age group (<5 years, 5-15 years, >15 years), gender, fever, study arm, and 2016 EA incidence.

|  | Unadjusted | | Adjusted | |
| --- | --- | --- | --- | --- |
|  | **OR (95%CI)** | **p-value** | **aOR (95%CI)** | **p-value** |
| Etramp5.Ag1 sero-positive | 1.97 (1.25 – 3.10) | 0.004 | 1.61 (1.90 – 2.59) | 0.049 |
| Age group |  |  |  |  |
| <5 years (reference) | -- | -- | 1.00 | -- |
| 5-15 years | -- | -- | 0.84 (0.48 – 1.47) | 0.540 |
| >15 years | -- | -- | 1.25 (0.76 – 2.07) | 0.376 |
| Gender |  |  |  |  |
| Female (reference) | -- | -- | 1.00 | -- |
| Male | -- | -- | 0.68 (0.45 – 1.04) | 0.073 |
| Fever | -- | -- | 9.36 (3.41 – 25.67) | <0.001 |
| Study arm |  |  |  |  |
| RACD only (reference) | -- | -- | 1.00 | -- |
| rfMDA | -- | -- | 0.99 (0.50 – 1.99) | 0.986 |
| RAVC | -- | -- | 0.93 (0.43 – 2.02) | 0.848 |
| rfMDA plus RAVC | -- | -- | 0.12 (0.01 – 0.98) | 0.048 |
| EA baseline incidence 2016 | -- | -- | 1.01 (1.00 – 1.02) | 0.011 |

Figure S7. Sero-prevalence compared to qPCR prevalence and clinical incidence rate. Comparison of sero-prevalence vs qPCR prevalence (A) and sero-prevalence vs. clinical incidence (B) are shown on the scatter plot on the log-scale. Each data point represents a study cluster, with the point diameter indicating either qPCR sample size per cluster (>100, 50-100, or <50 individuals) or person years at risk (PYAR) per cluster (>300, 200-300, or <200) and 95%CI for prevalence or incidence represented by the horizontal and vertical lines. Clusters in the RACD only study arm are indicated by the darker points to highlight the range of baseline prevalence and incidence values in the control arms.

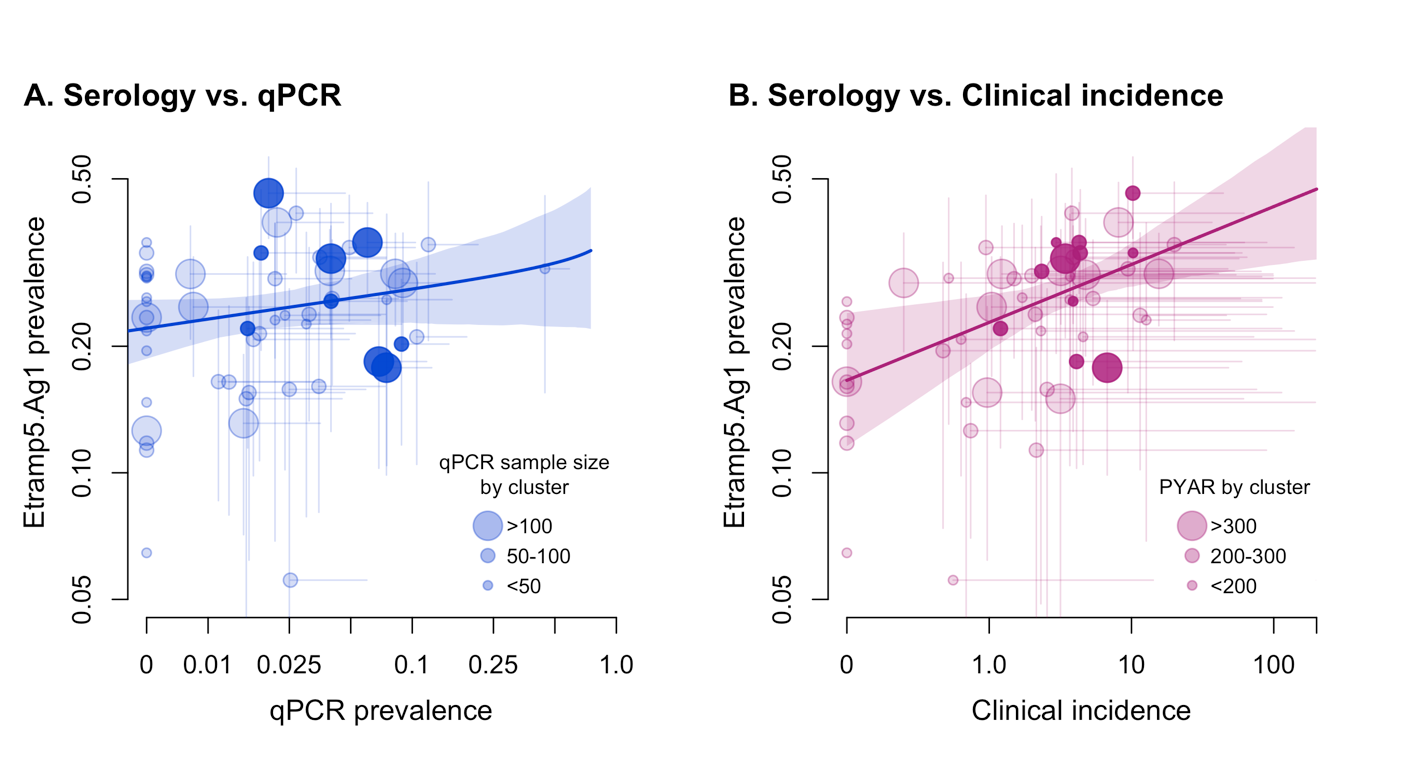

**Table S8. Relationship between cluster-level sero-prevalence, qPCR-prevalence, and clinical incidence.** Mean and 95% credible interval (CrI) of posterior from MCMC model fit shown for parameters ⍺_0_ (baseline log odds or log sero-prevalence), 𝛽_0_ (increase in log odds or log sero-prevalence), 𝛽_1,_ 𝛽_2_ (fixed effect of rfMDA, RAVC on rate of change in sero-prevalence per unit increase in qPCR prevalence or clinical incidence) and 𝛽_3_ (interaction effect of rfMDA and RAVC on rate of change in sero-prevalence per unit increase in qPCR prevalence or clinical incidence).

| Serology vs | qPCR (log odds scale) | Clinical incidence (log scale) |
| --- | --- | --- |
| Parameter | **Mean (95% CrI)** | |
| ⍺_0_ (intercept) | -0.71 (-1.12, -0.29) | -1.39 (-1.57, -1.22) |
| 𝛽_0_ (slope) | -0.94 (-1.03, -0.29) | 0.13 (0.04, 0.22) |
| 𝛽_1_ (rfMDA) | -0.17 (-0.52, 0.16) | -0.04 (-0.24, 0.16) |
| 𝛽_2_ (RAVC) | -0.09 (-0.42, 0.23) | -0.08 (-0.28, 0.11) |
| 𝛽_3_ (rfMDA:RAVC) | -0.28 (-0.79, 0.20) | -0.17 (-0.48, 0.13) |

**References**

1.	Burghaus, P. A. & Holder, A. A. Expression of the 19-kilodalton carboxy-terminal fragment of the Plasmodium falciparum merozoite surface protein-1 in Escherichia coli as a correctly folded protein. *Mol. Biochem. Parasitol.* **64**, 165–169 (1994).

2.	Collins, C. R. *et al.* Fine mapping of an epitope recognized by an invasion-inhibitory monoclonal antibody on the malaria vaccine candidate apical membrane antigen 1. *J. Biol. Chem.* **282**, 7431–41 (2007).

3.	Theisen, M., Vuust, J., Gottschau, A., Jepsen, S. & Høgh, B. Antigenicity and Immunogenicity of Recombinant Glutamate-Rich Protein of Plasmodium falciparum Expressed in Escherichia coli. **2**, 30–34 (1995).

4.	Richards, J. S. *et al.* Association between Naturally Acquired Antibodies to Erythrocyte‐Binding Antigens of *Plasmodium falciparum* and Protection from Malaria and High‐Density Parasitemia. *Clin. Infect. Dis.* **51**, e50–e60 (2010).

5.	Triglia, T. *et al.* Identification of proteins from Plasmodium falciparum that are homologous to reticulocyte binding proteins in Plasmodium vivax. *Infect. Immun.* **69**, 1084–92 (2001).

6.	Spielmann, T., Fergusen, D. J. P. & Beck, H.-P. etramps, a new Plasmodium falciparum gene family coding for developmentally regulated and highly charged membrane proteins located at the parasite-host cell interface. *Mol. Biol. Cell* **14**, 1529–44 (2003).

7.	Helb, D. A. *et al.* Novel serologic biomarkers provide accurate estimates of recent Plasmodium falciparum exposure for individuals and communities. *Proc. Natl. Acad. Sci. U. S. A.* **112**, E4438-47 (2015).
